# Proteomic aging clock predicts mortality and risk of common age-related diseases in diverse populations

**DOI:** 10.1101/2023.09.13.23295486

**Authors:** M. Austin Argentieri, Sihao Xiao, Derrick Bennett, Laura Winchester, Alejo J. Nevado-Holgado, Ashwag Albukhari, Pang Yao, Mohsen Mazidi, Jun Lv, Liming Li, Cassandra J. Adams, Robert Clarke, Najaf Amin, Zhengming Chen, Cornelia M. van Duijn

## Abstract

Circulating plasma proteins play key roles in human health and could be used to measure biological aging to predict risk of mortality, disease, and multimorbidity beyond chronological age. We developed a proteomic age clock using 1,459 plasma proteins (Olink Explore) in two prospective biobanks in the UK (n=45,117) and China (n=2,026) and explored its utility to predict incident risk of 26 major age-related diseases and all-cause mortality. We identified 226 proteins that accurately predicted chronological age (Pearson r=0.92). Individuals in the top versus bottom deciles of accelerated proteomic aging differed by approximately 10 years of biological aging. In the UK population, accelerated proteomic aging was associated with 25 aging phenotypes (e.g., telomere length, IGF-1, creatinine, cystatin C, hand grip strength, cognitive function, frailty index), 18 chronic diseases (e.g., diseases of the heart, liver, kidneys, lungs; diabetes; neurodegeneration; cancers), multimorbidity, and all-cause mortality. In the smaller Chinese population, accelerated proteomic aging was associated with ischemic heart disease, stroke, and all-cause mortality. Our results demonstrate that plasma proteins are a reliable instrument for prediction of multiple common diseases in diverse populations and can be used as a robust biochemical aging signature to improve early detection and management of common diseases.

## Main

Age is one of the most important risk factors for the development of most common diseases and causes of death.^1,2^ Aging involves a progressive loss of physiological integrity and function over time, which leads to an increased vulnerability to age-related diseases and death. Major chronic diseases such as ischemic heart disease (IHD), stroke, diabetes, liver and kidney disease, neurodegenerative disease, and some cancers all have varying rates of increasing risk with age.^2,3^ Importantly, there is significant variation across individuals in how quickly age-related diseases develop and how steeply mortality risk increases with age. How fast we age ultimately shapes the extent of middle and late life morbidity and disability, and determines whether premature mortality or longevity is achieved.^2^ The ability to quantify, and possibly intervene upon, rates of aging may therefore have important consequences for prevention of premature death by increasing population healthspan (years of disease-free life) and attenuating increasing rates of disease and multimorbidity.^4^

Chronological age is an imperfect surrogate measure of aging-related morbidity trajectories. Instead, aging-related biological changes can be estimated more precisely by using so-called biological aging clocks to capture the rate of age-related biological decline present more accurately in an individual. Some of the earliest and most successful biological aging clocks developed to date have used DNA methylation (DNAm).^5,6^ Loss of proteostasis is another primary hallmark of aging,^7^ and protein expression levels may provide a more direct mechanistic and functional insight into aging biology compared with DNAm.^6^ While several previous studies have systematically examined aging-related proteins (APs) and developed proteomic aging clocks,^8–11^ these studies have been constrained by smaller sample sizes (typically < 10k samples), lack of geographical diversity in their study populations, and inability to systematically evaluate associations between proteomic aging, biological markers of aging, aging-related physical and cognitive decline, and incidence of common diseases. To date, no studies have been reported that comprehensively assess the associations between accelerated biological aging (either DNAm or proteomic) and incidence of common diseases or major causes of death.

To address this gap in evidence, we explore the utility of using blood proteomic information to capture biological aging in a geographically diverse sample of participants from the UK Biobank (UKB) and China Kadoorie Biobank (CKB). We systematically assess the influence of accelerated proteomic aging (ProtAgeAccel; measured as the difference between protein predicted age and chronological age) on 27 aging-related phenotypes related to physical and cognitive decline, all-cause mortality, and 26 common age-related diseases that are either in the top 20 causes of death in the UKB or are highly prevalent in aging populations (rheumatoid arthritis, macular degeneration, osteoarthritis, osteoporosis).

### Proteomic aging clock

A schematic representation of our study design is shown in Fig. 1. Plasma proteomic profiling was conducted on 45,117 UKB participants (54% women, 46% men) between the ages of 39-71 years at baseline (mean 57.4 ± 8.2 years), as well as 2,026 CKB participants (62% women, 38% men) aged 30-78 years at baseline (mean 51.3 ± 10.5 years). Age distributions across cohorts are shown in Fig 2a. Across 11-16 years of follow-up in the UKB and 11-14 years of follow-up in the CKB, there were 4,784 (10.6%) and 182 deaths (9%) deaths, respectively (Fig. 2b). The prevalence and incidence rates of all 14 common diseases studied in the UKB are shown in Fig. 2c.

**Fig. 1.**
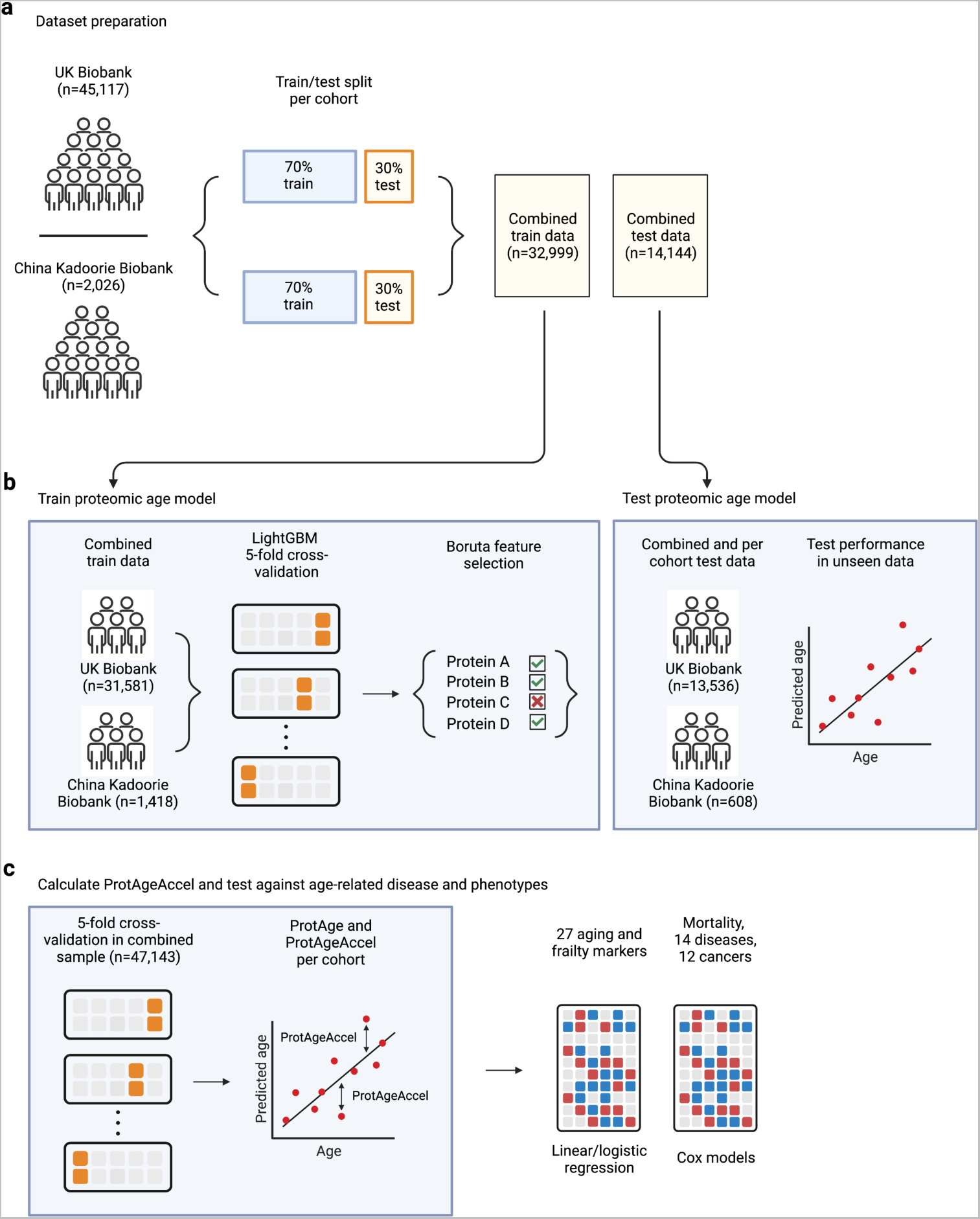
Overview of the study design. **a)** UK Biobank (UKB) and China Kadoorie Biobank (CKB) participants were both split into 70/30 training/test sets. **b)** The training sets from each cohort were combined and used to train the proteomic aging clock model using LightGBM and Boruta feature selection. The trained model was then tested on the holdout test set from each cohort, as well as the combined test sets. **c)** Protein predicted age (ProtAge) was then calculated in the full sample of all cohorts using 5-fold cross-validation, with proteomic age acceleration (ProtAgeAccel) calculated as the difference between ProtAge and chronological age in each cohort. ProtAgeAccel was tested in relation to a comprehensive panel of biological aging markers and measure of frailty and physical/cognitive decline, as well as mortality, 14 common diseases, and 12 cancers. Most association analyses were carried out in the UKB only, due to smaller sample size in the CKB.

**Fig. 2.**
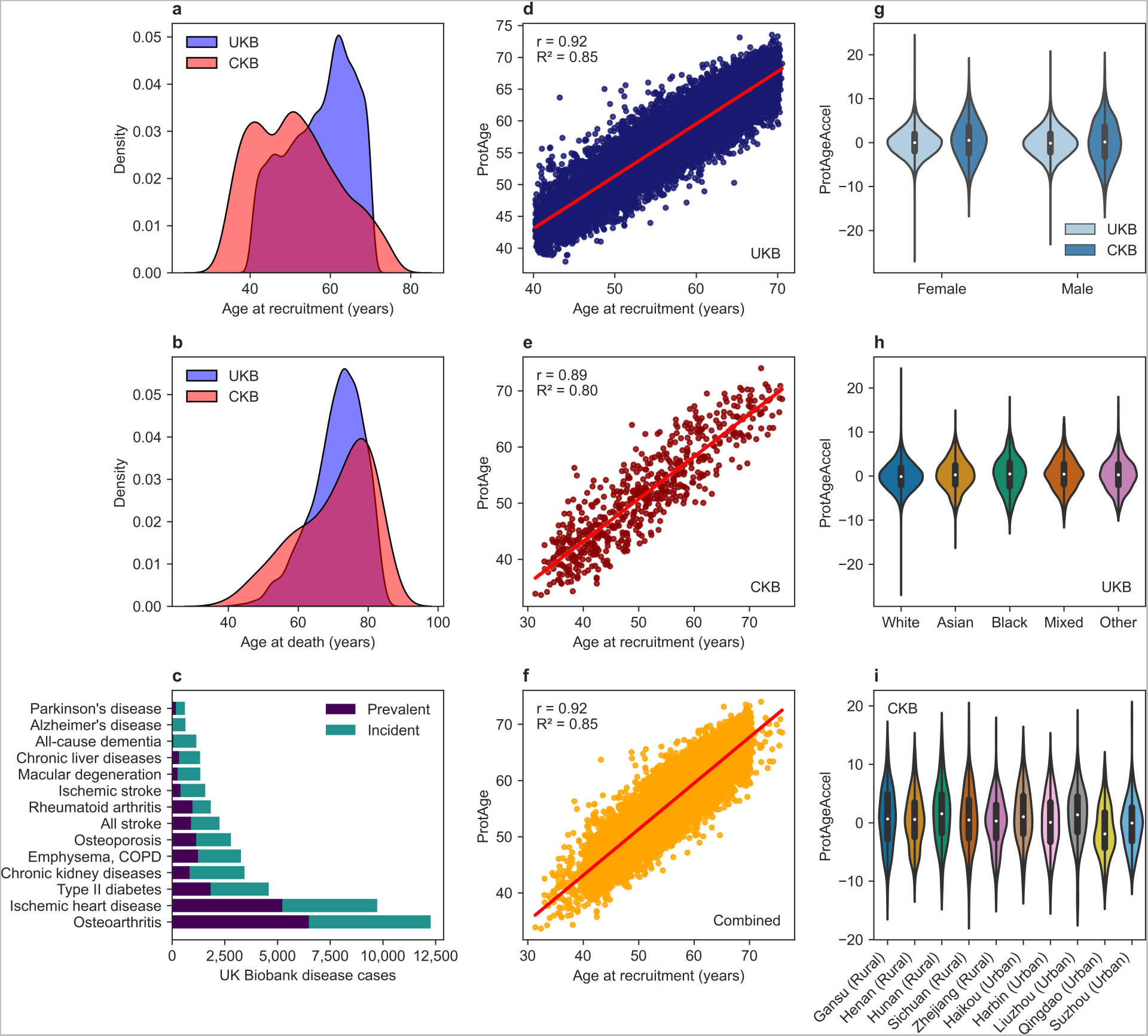
Proteomic aging clock performance across cohorts. **a)** Density plot of age at recruitment in the UK Biobank (UKB) and China Kadoorie Biobank (CKB). **b)** Density plot of age at death in the UKB (4,784 deaths [10.6%]) and CKB (182 deaths [9%]). **c)** Counts of prevalent and incident cases of all common diseases studied in the UKB sample (n=45,117). **d)** Performance of the trained proteomic aging model in the UKB holdout test set (n=13,536). **e)** Performance of the trained proteomic aging model in the CKB holdout test set (n=608). **f)** Performance of the trained proteomic aging model in the combined UKB and CKB holdout test sets (n=14,144). **g)** Sex distributions of ProtAgeAccel in the UKB and CKB. **h)** Distributions of ProtAgeAccel according to self-reported ethnicity in the UKB. **i)** Distributions of ProtAgeAccel according to geographic region of residence in the CKB. Correlation coefficients shown in **d-f** are Pearson correlation coefficients. Violin plots in **g-i** show both the median (white dot) and interquartile range. COPD: chronic obstructive pulmonary disease, ProtAge: protein predicted age, ProtAgeAccel: proteomic age acceleration (in years).

We randomly split both the UKB and CKB cohorts into 70% training and 30% test sets, and within the combined training sets (n=32,999) we trained a single LightGBM machine learning model including normalized expression of all 1,459 proteins to predict chronological age. Boruta feature selection identified a subset of 226 APs (Supplementary File SF1). Protein predicted age (ProtAge) from our models using the subset of 226 APs explained a high degree of variation in chronological age (R^2^: 0.78-0.85) and was strongly correlated with chronological age (Pearson r: 0.89-0.92) in both the UKB (n=13,536) and CKB (n=608) holdout test sets, as well as the combined test sets (n=14,144) (Fig. 2d-f). Using the subset of 226 APs did not result in an appreciable decline in model performance compared with the predictive model using all 1,459 proteins (R^2^ in combined test data: 0.856). ProtAgeAccel showed similar distributions in both datasets, and showed similar distributions in women and men in both cohorts, across self-reported ethnicities in the UKB, and across geographical regions in the CKB (Fig. 2g-i). While our ProtAge clock was developed using a combined UKB and CKB sample, ProtAgeAccel was tested in relation to diseases and aging phenotypes in each cohort separately, with most association analyses carried out in just the UKB because of the small sample size in the CKB.

The reliability of each AP’s association with age was tested using repeat protein expression measurements for subsets of UKB participants with repeat protein expression data available. Specifically, repeat protein measurements were available for a subset of n=114 participants at the imaging study visit (2014+) and n=108 participants at the repeat imaging visit (2019+). Associations of each of the 226 APs with age were tested during the baseline visit in addition to the two follow-up visits using linear regression. The beta coefficients across all three time points were strongly correlated with each other (Pearson r = 0.81-0.84), confirming the stability of associations between the 226 APs and age across repeat visits spanning at least 9-13 years (Extended Data Fig. 1).

### Biological functions and interaction networks of proteomic aging

The 226 proteins we identified as predictive of age provide a preliminary map of APs from the Olink Explore 1536 assay in the UKB and CKB. Testing for functional enrichments among the 226 APs according to Gene Ontology (GO) biological processes and molecular function, as well as Kyoto Encyclopedia of Genes and Genomes (KEGG) and Reactome, identified many statistically significant enrichments (Fig. 3a-d). The strongest enrichments (top 20 pathways for each database according to FDR) can be organized thematically into several larger categories of biological functions: (i) cellular signaling and regulation, (ii) development and growth, (iii) immune responses and inflammation, (iv) cell adhesion and extracellular matrix (ECM) interactions, (v) metabolic processes and regulation, (vi) apoptosis and cell death, and (vii) disease-related processes.

**Fig. 3.**
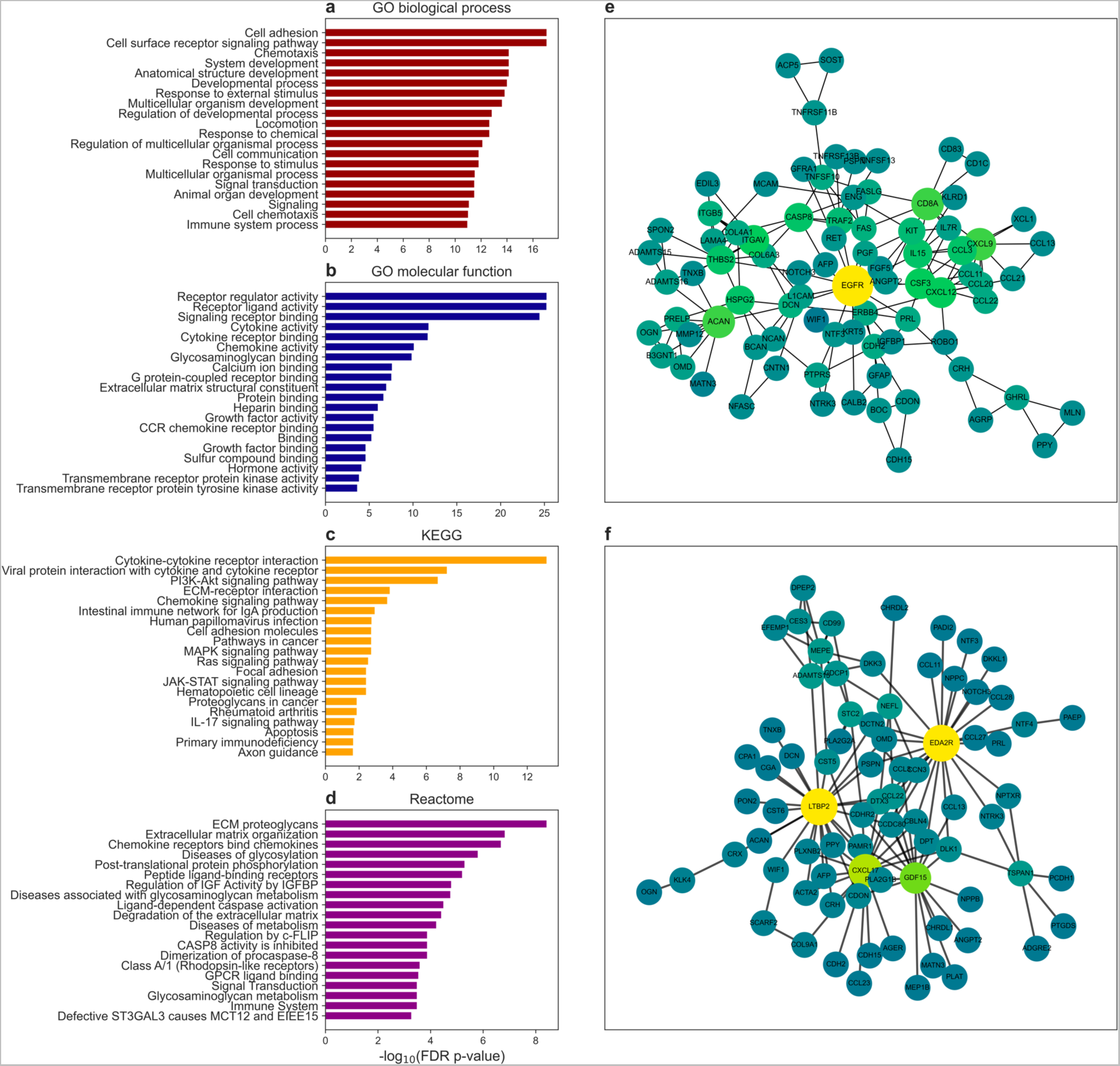
Protein-protein interaction network and pathway enrichments of ProtAge. The top 20 most FDR significant pathway enrichments are shown for the 226 ProtAge APs according to: **a)** GO biological process, **b)** GO molecular function, **c)** KEGG, and **d)** Reactome. **e)** Protein-protein interaction (PPI) network from the STRING database for a highly interconnected subnetwork of 83 proteins in ProtAge with at least 2 node connections. **f)** PPI network for a highly interconnected subnetwork of 78 ProtAge APs using SHAP interaction values from the trained ProtAge model. In **e** and **f**, nodes are colored by number of connections (degrees). Darker nodes have fewer connections, whereas light nodes have more connections. APs: aging-related proteins, ECM: extracellular matrix, FDR: false discovery rate, GO: Gene Ontology, KEGG: Kyoto Encyclopedia of Genes and Genomes.

Overall, these 226 APs showed a large number of protein-protein interactions (PPIs). Among these 226 APs, we identified a highly interconnected subnetwork of proteins consisting of 83 proteins with at least 2 node connections in a PPI network using co-expression information from the STRING database (Fig. 3e). The key proteins with the greatest numbers of connections to other proteins were EGFR (involved in cancer drug resistance, brain structure, and platelet count), ACAN (an integrin protein involved in body mass index [BMI], blood pressure, lipid metabolism, and renal agenesis), ITGAV (an integrin protein implicated in body height, handedness, dyslexia, and albumin/creatinine metabolism), CXCL9 (implicated in T-cell function and inflammation), and CD8A (a CD8 antigen implicated in the innate immune system).

We also used SHAP interaction values from our trained ProtAge model to calculate a second PPI network that represents the interactions of proteins together in the model to predict age (Fig. 3f). The key proteins with the largest numbers of connections to other proteins according to SHAP interaction values were EDA2R (involved in the NF-κB and innate immune pathways and implicated in baldness, estradiol, testosterone and HDL metabolism), LTPB2 (a protein involved in BMI, blood pressure, neuroticism and anxiety, glaucoma and retina pathology, lung function and mortality), CXCL17 (a chemokine interacting with CXCL9, that plays a role in tumor genesis, antimicrobial defense through monocytes, macrophages, and dendritic cells), and GDF15 (implicated in BMI, liver function, systemic lupus erythematosus, and COVID-19). This second network provides a visual representation of how the most influential APs in our ProtAge model interacted in the prediction of age, whereas the STRING-based PPI network represents interactions related to known biological connections between proteins. Overall, we found quite distinct results when using a data driven approach to modelling PPIs using interactions from our machine learning models versus using experimental biological knowledge from the STRING database.

### Proteomic aging predicts frailty and biological decline even before disease onset

To understand how accelerated proteomic aging may influence aging-related physiological and cognitive decline, we tested associations in the UKB between ProtAgeAccel and: (i) a comprehensive frailty index (see Methods); (ii) 16 individual measures of physical (e.g., slow walking pace, grip strength) and cognitive (reaction time, fluid intelligence) decline, and (iii) 10 measures of biological aging and blood biochemistry (e.g., telomere length, IGF-1, creatinine). After adjustment for chronological age, sex, and major sociodemographic and lifestyle confounders, ProtAgeAccel aging was significantly associated with all measures tested except for alanine aminotransferase (ALT) and total bilirubin (Fig. 4a-b).

**Fig. 4.**
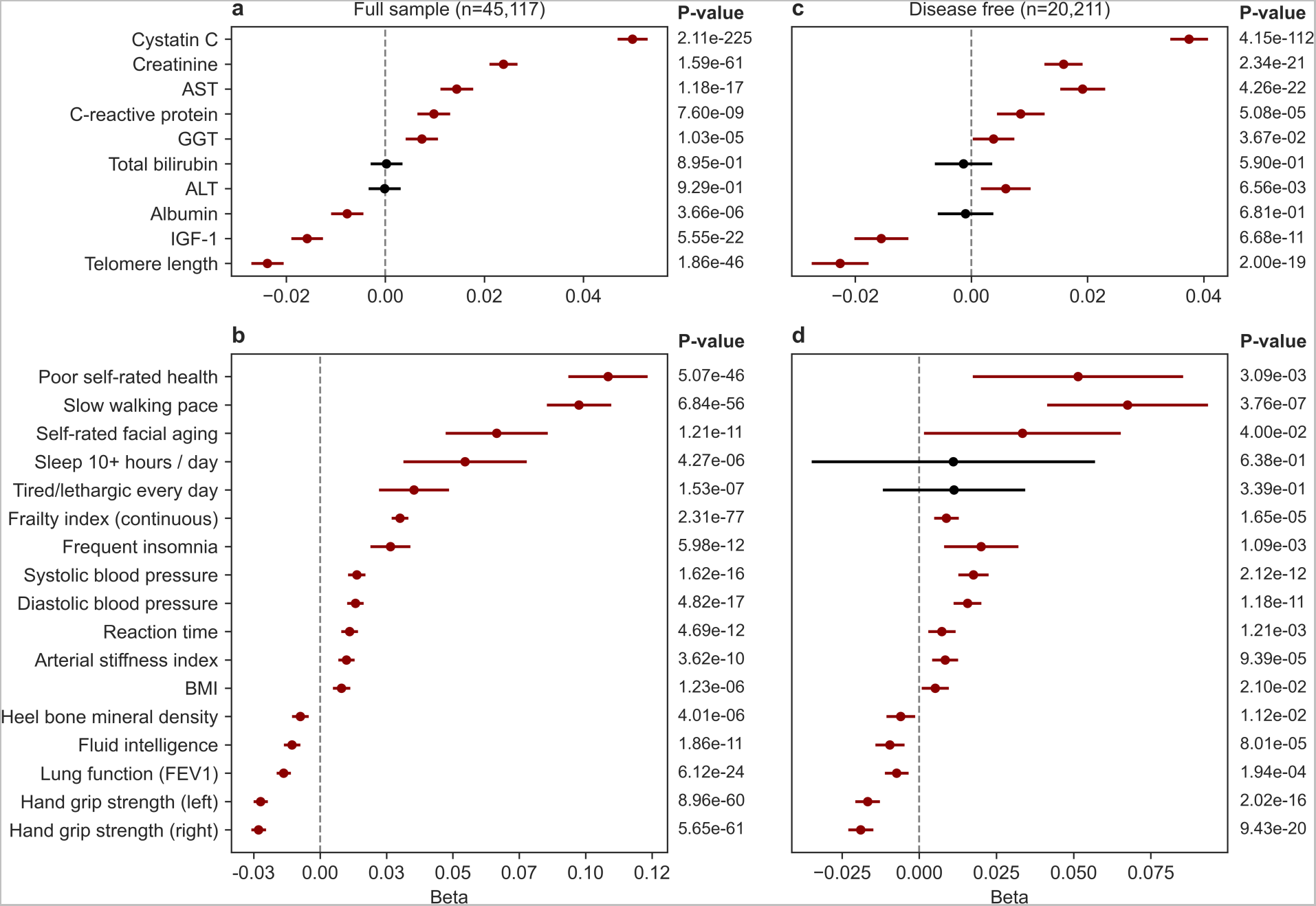
ProtAgeAccel is associated with age-related biological, physical, and cognitive decline. **a)** Associations between ProtAgeAccel and biological aging mechanisms in the full UKB sample (n=45,117). **b)** Associations between ProtAgeAccel and measures of physiological and cognitive (reaction time, fluid intelligence) decline in the full UKB sample (n=45,117). **c)** Associations between ProtAgeAccel and biological aging mechanisms in the subsample of UKB participants with no lifetime diagnosis of any of the 26 diseases studied (n=20,211). **d)** Associations between ProtAgeAccel and measures of physiological and cognitive decline in the subsample of UKB participants with no lifetime diagnosis of any of the 26 diseases studied (n=20,211). All models used linear or logistic regression and were adjusted for age, sex, Townsend deprivation index, recruitment centre, ethnicity, IPAQ activity group, and smoking status. ALT: alanine aminotransferase, AST: aspartate aminotransferase, BMI: body mass index, FEV1: forced expiratory volume in 1 second, GGT: Gamma-glutamyl Transferase, IGF-1: insulin-like growth factor 1, ProtAgeAccel: proteomic age acceleration (in years).

Among biological aging mechanisms tested (Fig. 4a), increasing ProtAgeAccel was associated with increasing levels of Cystatin C, Creatinine, aspartate aminotransferase (AST), C-reactive protein, and gamma-glutamyl transferase (GGT); and was associated with decreased levels of albumin, IGF-1, and telomere length.

Among physical measures tested (Fig. 4b), increasing ProtAgeAccel was associated with poor self-rated health, slow walking pace, self-rating one’s face as older than average, sleeping 10+ hours per day, feeling tired every day, and having frequent insomnia. It was also associated with increased values of a comprehensive frailty index, as well as increased blood pressure, longer (slower) reaction time, greater arterial stiffness, and greater BMI. Moreover, ProtAgeAccel was associated with decreased bone mineral density, fluid intelligence, lung function, and hand grip strength.

Associations between ProtAgeAccel and all aging phenotype measures were also tested in the subset of UKB participants with no lifetime diagnoses of any of the 26 diseases studied (n=20,211). Among these participants, we found that ProtAgeAccel remained significantly associated with nearly all markers tested (Fig. 4c-d). Of the physical measurements, only sleeping for 10+ hours/day, and feeling tired every day were no longer significantly associated with ProtAgeAccel. Interestingly, albumin was no longer significant in this subset of participants, despite being significantly inversely associated with ProtAgeAccel in the full dataset. ALT became significant while not being significant in the full dataset. All other biochemical markers remained significantly associated with ProtAgeAccel in the disease-free subsample.

### Proteomic age acceleration is a strong predictor of common diseases

UKB participants in the top, median, and bottom deciles of ProtAgeAccel showed dramatically divergent age-specific incidence rates of all-cause mortality and the 14 common non-cancer diseases studied (Fig. 5a). The average years of biological age acceleration among the top 10% of ProtAgeAccel was 5.6 years and the average among the bottom 10% was −5.4 years, resulting in an average difference of approximately 10 years in biological aging between the top and bottom deciles of ProtAgeAccel. For those aged 65 years at recruitment, the highest cumulative incident rates (equivalent to absolute risk) across the study follow-up period of 11-16 years for the top decile of ProtAgeAccel were observed for osteoarthritis (76.65%), all-cause mortality (60.02%), chronic kidney diseases (CKD; 53.27%), IHD (47.60%) and type II diabetes (47.49%). Neurodegenerative diseases (Parkinson’s disease, all-cause dementia, Alzheimer’s disease [AD]) and ischemic stroke all showed cumulative incident rates below 1% in the bottom decile of ProtAgeAccel across all ages. Diseases with the greatest difference in cumulative incidence rate between the top and bottom deciles of ProtAgeAccel among those aged 65 years at recruitment included osteoarthritis (Δ 66.02%), all-cause mortality (Δ 56.03%), CKD (Δ 51.17%), type II diabetes (Δ 43.74%), and IHD (Δ 42.26%). Cumulative incidence risk trajectories according to these deciles of ProtAgeAccel looked similar when plotted separately in women and men (Extended Data Fig. 2–3). For all diseases, cumulative incidence rates in the UKB across follow-up for those aged 50-65 years at recruitment are shown in Table S1.

**Fig. 5.**
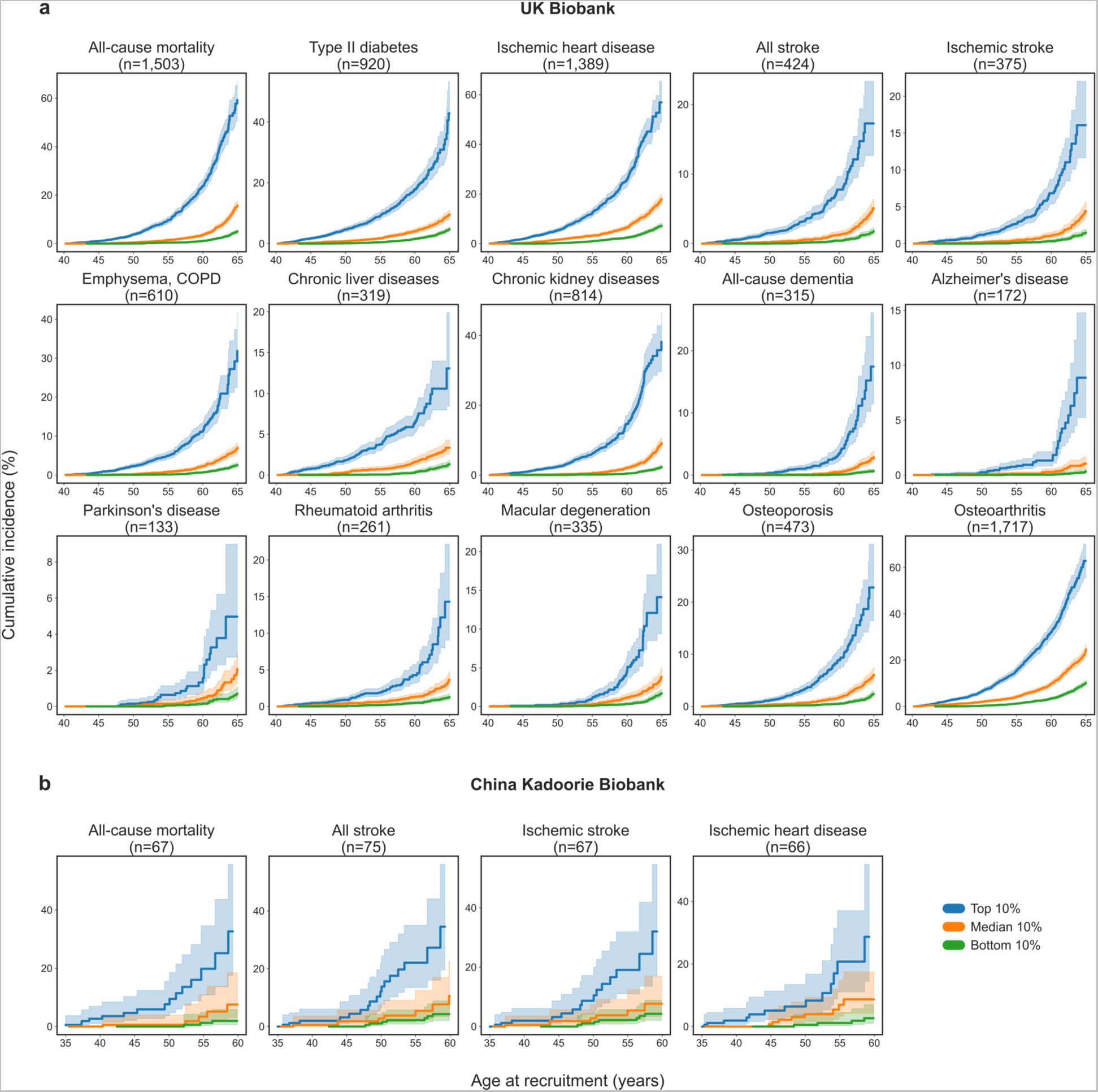
ProtAgeAccel lead to strongly diverging age-specific mortality and disease risk trajectories in the UKB and CKB. Cumulative incidence plots for the top, median, and bottom deciles of ProtAgeAccel in **a)** UK Biobank (UKB; n=45,117) and **b)** China Kadoorie Biobank (CKB; n=2,026) samples. Number of incident cases are shown for each disease – these numbers reflect the total number of incident cases present only among those in the 3 deciles shown, not the full dataset. Incidence rates are shown for the subsequent 11-16 years (UKB) or 11-14 years (CKB) of follow-up after recruitment for each given age at recruitment (e.g., the cumulative incidence rate shown at age 65 in **a)** is the rate of incident cases in the 11-16 years of follow up for those aged 65 at recruitment). All plots show 95% confidence intervals in lighter shading. Diseases shown here for the CKB are those with greater than 50 cases across the three deciles of ProtAgeAccel. Plots for all diseases in the CKB are shown in Fig. S1. ProtAgeAccel: proteomic age acceleration (in years).

In the CKB, we also calculated cumulative incidence rates according to deciles of ProtAgeAccel, although the smaller sample size restricted the number of diseases that could be reliably analyzed. Plots for incident outcomes with greater than 50 incident cases across the 3 deciles of ProtAgeAccel (mortality, IHD, all stroke, ischemic stroke) are shown in Fig. 5b. For those aged 60 years at recruitment, the cumulative incidence rates across the study follow-up period for the top decile of ProtAgeAccel were 47.64% (all stroke), 45.61% (ischemic stroke), 32.65% (all-cause mortality), and 28.69% (IHD). Cumulative incidence plots for all diseases with at least one incident case in each of the 3 deciles of ProtAgeAccel are shown in Fig. S1. Cumulative incidence rates in the CKB across follow up for those aged 36-65 years at recruitment are shown in Table S2.

We further tested whether associations of ProtAgeAccel with mortality and the 14 common diseases were robust to adjustment for chronological age, sex, smoking, physical activity, sociodemographics, and clinical risk factors using multivariable Cox proportional hazards models. ProtAgeAccel showed a significant association with mortality and all disease outcomes across all models tested in the UKB (Fig. 6). In the fully adjusted model that also included covariates for BMI and prevalent hypertension (Model 3), ProtAgeAccel showed the largest effect size for AD (HR: 1.18; 95% CI: 1.15-1.22), followed by all-cause dementia (HR: 1.15; 95% CI: 1.12-1.18) and CKD (HR: 1.13; 95% CI: 1.11-1.14). Hazard ratios for each outcome are given per year increase of ProtAgeAccel, therefore those in the top 10% of ProtAgeAccel have on average a 2.6 times higher risk of AD than those with no acceleration (HR of 1.18^5.6^ = 2.6), and a 6.5 times higher risk of AD (HR of 1.18(5.6+[–5.4])) than those in the bottom 10% of biological age acceleration. For CKD, the increases in risk are 1.9 (top 10% vs. 0) and 3.7 times (top 10% vs. bottom 10%), and for mortality the increases in risk are 1.8 (top 10% vs. 0) and 3.1 times (top 10% vs. bottom 10%).

**Fig. 6.**
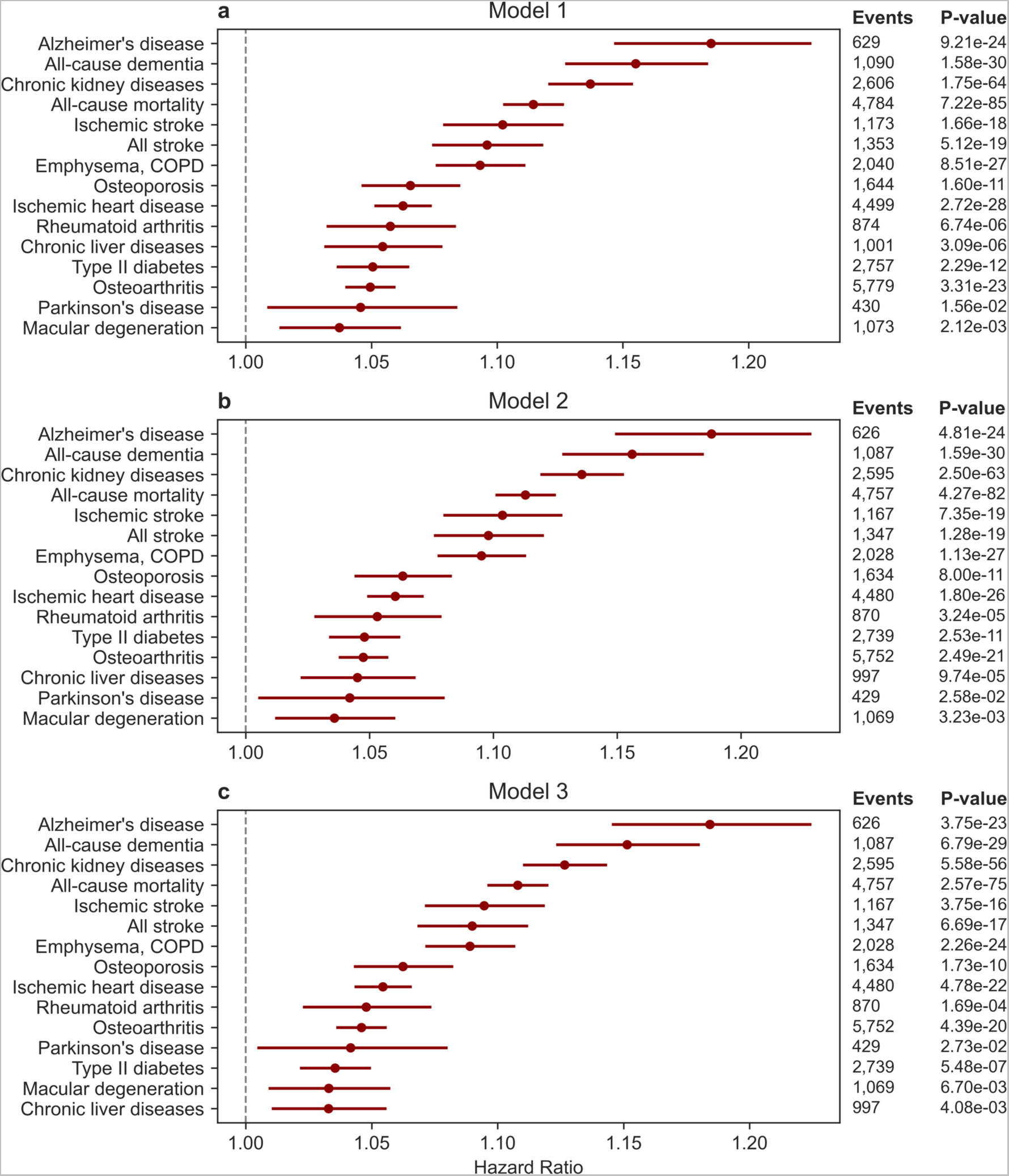
Effect size of ProtAgeAccel on mortality and common diseases is largely invariant to covariate adjustment. Associations between ProtAgeAccel and mortality or diseases in Cox proportional hazards models with increasing levels of covariate adjustment. All models were run in the UK Biobank (UKB; n=45,117). **a)**. Model 1 is adjusted for age and sex. **b)** Model 2 is adjusted for age, sex, Townsend deprivation index, recruitment centre, IPAQ activity group, and smoking status. **c)** Model 3 is adjusted for age, sex, Townsend deprivation index, recruitment centre, IPAQ activity group, smoking status, BMI, and prevalent hypertension. ProtAgeAccel: proteomic age acceleration (in years).

In the UKB, we also investigated the relationships between ProtAgeAccel and incident cancer diagnoses (Fig. 7), but associations from Cox models were less robust to adjustment for age and other confounders. ProtAgeAccel was associated with only four cancers (esophageal, liver, lung, non-Hodgkin lymphoma) after adjustment for age, sex, sociodemographics, and lifestyle factors (Extended Data Fig. 4).

**Fig. 7.**
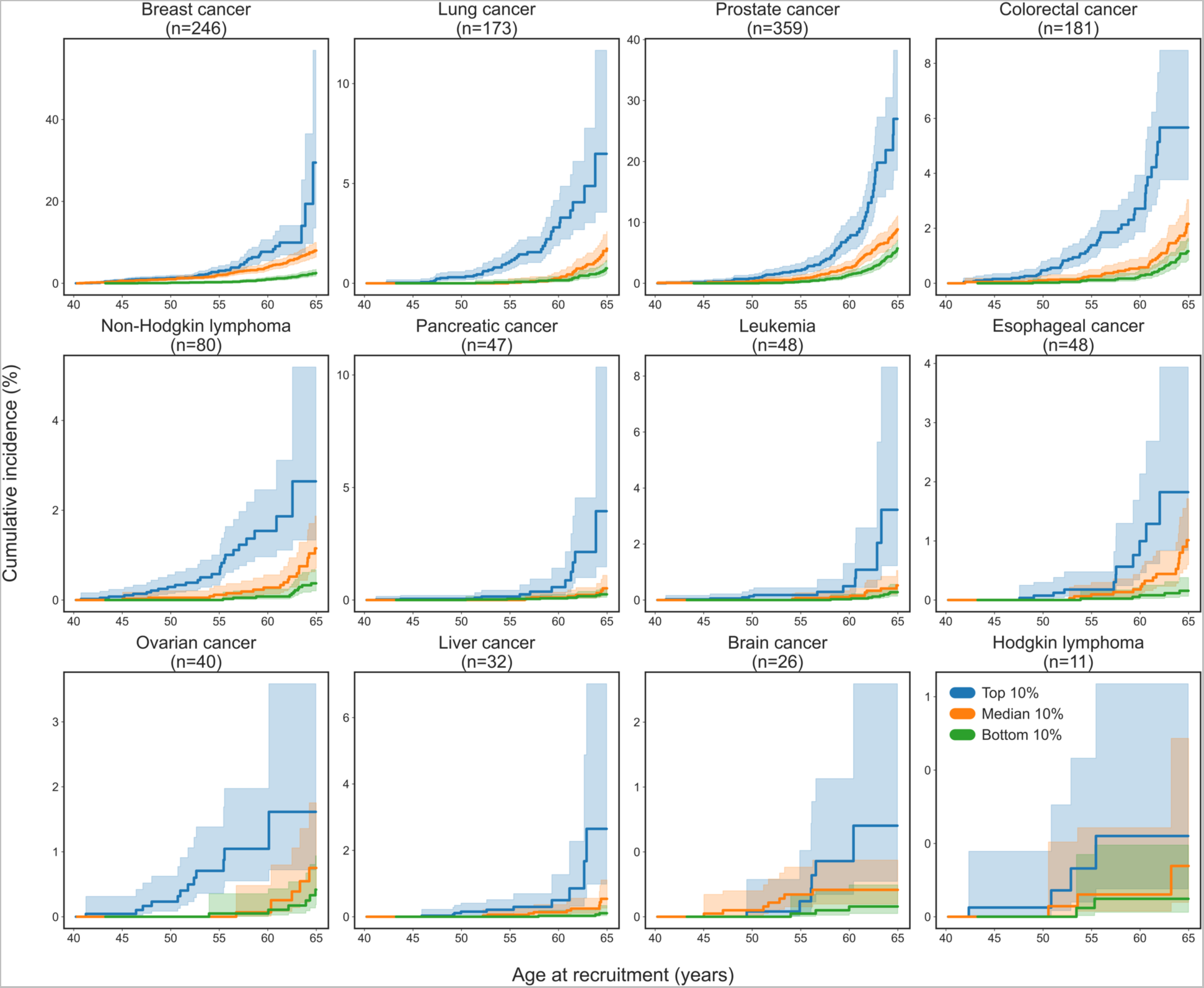
ProtAgeAccel and age-specific cancer risk trajectories in the UKB. Cumulative incidence plots for the top, median, and bottom deciles of ProtAgeAccel in the UK Biobank (UKB; n=45,117). Number of incident cases are shown for each cancer – these numbers reflect the total number of incident cases present only among those in the 3 deciles shown, not the full dataset. Incidence rates are for the 11-16 years after recruitment. Incidence rates are shown for the subsequent 11-16 years of follow up after recruitment for each given age at recruitment (e.g., the cumulative incidence rate shown at age 65 is the rate of incident cases in the 11-16 years of follow for those aged 65 at recruitment). All plots show 95% confidence intervals in lighter shading. Certain plots show 0 twice on the y-axis because these represent decimal values < 0.5 and the y-axis values are rounded to a single digit. ProtAgeAccel: proteomic age acceleration (in years).

Although the analyses described above were adjusted for smoking status, we conducted further sensitivity analyses testing associations of ProtAgeAccel with all-cause mortality and all 14 non-cancer common diseases in never smokers. Among never smokers, ProtAgeAccel remained significantly associated with all outcomes except Parkinson’s disease (Extended Data Fig. 5). In a similar sensitivity analysis restricted to those within a normal weight range (BMI ≥ 18.5 & BMI < 25), ProtAgeAccel remained significantly associated with all outcomes except Parkinson’s disease and macular degeneration (Extended Data Fig. 6). These results indicate that ProtAgeAccel picks up a significant biological aging signature that is independent of smoking and extreme BMI profiles.

### Years of ProtAgeAccel increase linearly with multimorbidity

We also investigated whether ProtAgeAccel is an informative marker of multimorbidity status in the UKB. We defined multimorbidity as lifetime diagnosis of any of the 26 diseases included in our study, and categorized participants according to having 0, 1, 2, 3, or 4+ lifetime diagnoses. We found that average years of ProtAgeAccel increased linearly with increasing number of comorbid conditions (Fig. 8a-b). We also found that this effect was more pronounced for younger participants at recruitment (aged 40-50 years; Fig. 8a), among whom presence of disease was less common (Fig. 8c). On average, those aged 40-50 years at recruitment with 4+ comorbid conditions had approximately 1.8 greater years of ProtAgeAccel compared to those with no lifetime diagnoses (Fig. 8a), whereas those aged 51-65 years at recruitment with 4+ comorbid conditions had approximately 0.9 greater years of ProtAgeAccel compared to those with no lifetime diagnoses (Fig. 8b). Average ProtAgeAccel also increased linearly with self-reported health, with those reporting excellent health showing and average −0.2 years of ProtAgeAccel (indicating that they are younger biologically than chronologically) and those with poor self-reported health showing and average 0.9 years of ProtAgeAccel (Fig. 7d).

**Fig. 8.**
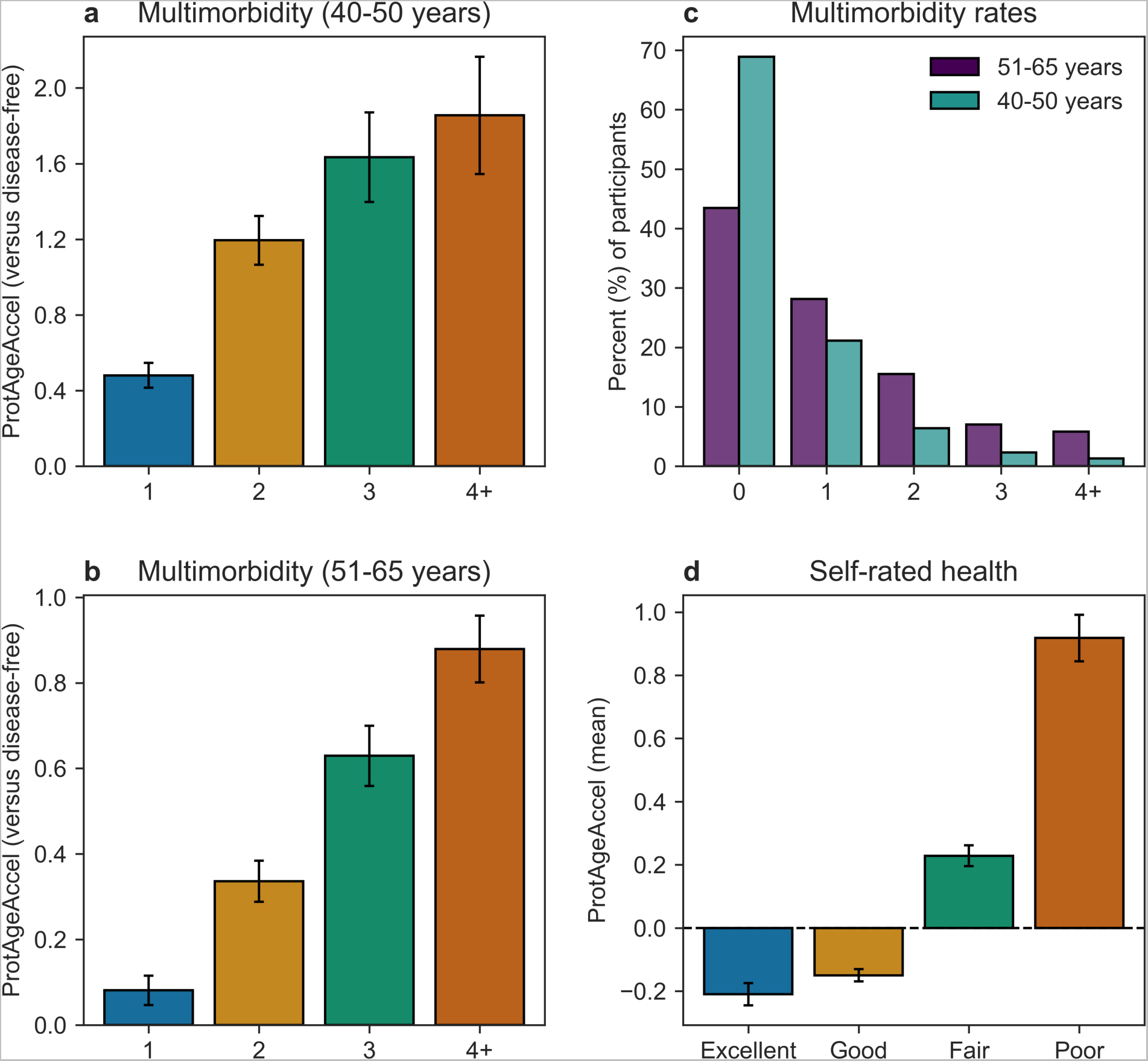
ProtAgeAccel increases linearly with increasing disease multimorbidity. **a)** Average years of ProtAgeAccel in those with 1 disease diagnosis or 2, 3, 4+ comorbid conditions compared with average ProtAgeAccel in those with no diagnoses among UK Biobank (UKB) participants 40-50 years old at recruitment. **b)** Average years of ProtAgeAccel in UKB participants with 1 disease diagnosis or 2, 3, 4+ comorbid conditions compared with average ProtAgeAccel in those with no diagnoses aged 51-65 years old at recruitment. **c)** Percentages of the UKB population with 0, 1, 2, 3, and 4+ lifetime disease diagnoses. **d)** Average years of ProtAgeAccel according to levels of self-rated health in the UKB. In **a)** and **b)**, values on the y-axis represent the average years of ProtAgeAccel for each group compared with the average in those with no diagnoses (calculated as the difference in average ProtAgeAccel between the two groups). Multimorbidity is defined as the number of lifetime diagnoses of any of the 26 diseases analyzed in this study. In **a**, **b**, and **d**, error bars are shown as the standard error of the mean. ProtAgeAccel: proteomic age acceleration (in years).

## Discussion

In the analyses presented here using data from both UK and Chinese populations, we demonstrate that ProtAgeAccel is strongly associated with a large and diverse range of markers related to physical frailty and cognitive decline, as well as measures of biological aging (e.g., telomeres, IGF-1, creatinine, cystatin C). While previous research has shown that DNAm aging is not related to telomere length,^12^ we show that proteomic aging is strongly inversely associated with telomere length – a key cellular hallmark of aging.^7^ Our study provides important new evidence that proteomic aging is a common feature underlying most aging-related frailty, cognitive decline, and biological decline. Importantly, our study also provides the most comprehensive evidence to date cataloguing the incident disease outcomes that are associated with proteomic aging, and provides the first systematic evidence within a single study that biological aging is a common signature underlying all common adult diseases. We find that ProtAgeAccel is a robust predictor of mortality and many of the most common causes of death in the UK, including all 14 common diseases tested and four cancers (esophageal, liver, lung, non-Hodgkin lymphoma).

Recently, there has been growing interest in using plasma proteins to study biological aging and create proteomic aging clocks. We specifically compared our findings to three of the largest published studies: (1) a systematic review of studies (n=32) reporting protein associations with age (Johnson et al. 2020^9^), in which the authors developed a proteomic age clock using 85 proteins associated with age in at least three previous studies and validated it in the INTERVAL cohort (n=3,301); (2) a recent study that identified 273 APs across several cohorts (Coenen et al. 2023^8^) (n=37,650); and (3) a clock consisting of 373 APs developed in the INTERVAL and LonGenity cohorts (Lehallier et al. 2019^10^) (n=4,263). To our knowledge, neither these nor any previous study has directly tested associations between proteomic aging and disease or multimorbidity in the comprehensive manner described here.

Furthermore, for outcomes that were measured in these previous proteomic clock analyses, our ProtAge clock shows improved performance. For the Lehallier et al. 2019 clock, p-values (but not betas) for associations with physical and cognitive frailty measures were reported. For similar traits, our ProtAgeAccel measure shows remarkably stronger p-values (e.g., hand grip strength: −log_10_ p-value of 43.6 in our study vs 3.8 in Lehallier et al. 2019), although this may be explained by a much larger sample in our analysis (n=45,117 vs. n=4,263). Coenen et al. 2023 report associations of their aging clock acceleration measure with select blood clinical markers. Their clock only showed marginally significant partial correlations with blood uric acid and magnesium, but non-significant correlations with the other 57 blood markers tested, including albumin, creatinine, GGT, and C-reactive protein, all of which were strongly associated with ProtAgeAccel in our study. The Johnson et al. clock was not tested against mortality, morbidity, or any frailty or cognitive measures. It is unclear whether lack of greater overlap between ProtAge APs and those from these previous papers is due to differences between measurement on the Olink and SOMAscan platforms. Previous research comparing SOMAscan and Olink platforms directly demonstrated substantial discrepancies in protein-phenotype associations and protein quantitative trait loci (pQTL) mapping between the two platforms.^17^

Our ProtAge clock also provides a technical advantage over many state-of-the-art DNAm clocks. Although the so-called ‘first generation’ DNAm clocks (e.g., Horvath clock) were able to accurately capture the chronological passage of time, they do not effectively capture aging-related biological decline.^5,6^ Newer DNAm clocks were subsequently developed that are not trained on chronological age itself, but rather are trained to predict composite variables that are usually weighted combinations of age and other biological aging markers (e.g., albumin, creatinine, C-reactive protein).^13–15^ This has been key to developing DNAm clocks that are more predictive of mortality, morbidity, and frailty.^5^ In contrast, we demonstrate that a proteomic aging clock can be constructed by training on age itself and remain a strong predictor of mortality, disease, multimorbidity, and frailty without the need for creating more complicated biological aging phenotypes used to train initial models. Compared to DNAm-based measures, the proteome appears to be a more powerful measure of biological aging because it is a functional layer directly involved in biological decline that is much more proximal to disease itself.

Proteins selected by our model showed very little overlap with corresponding genes from leading DNAm clocks, including the Horvath clock,^16^ PhenoAge,^13^ and DunedinPACE^15^ (Extended Data Fig. 7a). Six corresponding genes from ProtAge overlapped with genes mapped to CpGs (by proximity) in the Horvath clock (*CCL27*, *DKK3*, *ENPP2*, *LAG3*, *LGALS1*, *MCAM*), and nine ProtAge genes overlapped with proximity genes mapped to PhenoAge (*ACP5, CALB1, CST6, CTSF, DPEP2, KLK10, KLK8, LHB, MATN3*). Only one ProtAge gene overlapped with proximity genes mapped to DunedinPACE (*TNXB*). While the proteins selected by our model showed somewhat greater overlap with those found in existing proteomic clocks, 149 ProtAge APs (66%) were not identified in any of these major previous studies on proteomic aging (Extended Data Fig. 7b). Despite representing a largely novel set of APs, ProtAge also includes 16 APs present in the Johnson et al., Coenen et al., and Lehallier et al. analyses (Extended Data Fig. 7b, Extended Data Table 1). Interestingly, none of these 16 proteins overlap with corresponding genes from any of the DNAm clocks, indicating that DNAm and proteomic clocks appear to converge on differing gene sets. The overlap between the 226 ProtAge APs and all previous studies described here is represented in Supplementary File SF1.

Finding common pathways and upstream targets across major common diseases will be key to developing preventative measures to curb globally increasing rates of multimorbidity.^4^ We show that ProtAgeAccel is a common biological signature underlying most of the major non-communicable causes of death in the UK, and that years of ProtAgeAccel increase linearly with increasing comorbid conditions. Importantly, we also show that ProtAgeAccel is associated with nearly all measures of frailty and physical, cognitive, and biological decline even among those who have no lifetime diagnoses of any of the 26 common diseases studied. This indicates that proteomic age acceleration is a biological signature of frailty and disease susceptibility that can be measured and detected before the onset of disease itself. We posit that studying accelerated proteomic aging is a strategic starting point for understanding the common pathways driving disease multimorbidity. We find that ProtAge APs are significantly enriched for cell signaling, immune response, metabolic regulation, and inflammation pathways, which converges with results from previous studies of proteomic aging.^11^

Our functional enrichment analyses of ProtAge APs identified several cancer-related pathways, and our Cox regression results showed consistent associations between ProtAgeAccel and 4 cancers (lung, liver, esophageal, non-Hodgkin lymphoma). The reasons why the remaining cancers tested did not associate with ProtAgeAccel remain unclear. Some had extremely few cases, which likely influenced power to detect associations. For other cancers with adequate sample size (e.g., prostate cancer), it may be that the Olink panel we used doesn’t contain the relevant proteins for that cancer or that there is not a reliable signature in blood.

The lack of associations between ProtAgeAccel and ALT and total bilirubin are interesting and warrant further research. One plausible explanation is that some liver enzymes tend to reflect more acute liver injury,^18^ which may not necessarily reflect long term aging-related liver function decline or inflammation. In contrast, albumin is more reflective of liver function and productivity,^18^ as well as broader nutritional status, which may be more reflective of aging-related decline. Our functional analyses showed that the 226 ProtAge APs are significantly enriched for proteins involved in signaling of MAPK, JAK-STAT, PI3K-Akt, and IGF-1, all of which show altered expression in senescent hepatocytes (linked to chronic liver disease).^19^

Our study was conducted in the largest sample size to date for a proteomic aging clock, included a geographically diverse training population, and included a wide range of phenotypes and diseases. Furthermore, our use of LightGBM brings significant improvements over existing DNAm and proteomic clock approaches, since our model allows for non-linearities and accounts for interactions between all proteins. Our study also has several limitations to note. First, our model only uses the Olink Explore 1536 assay currently available in the UKB, and therefore did not capture all proteins covered in the larger Olink Explore + Expansion panel or larger SOMAscan panels. Second, we have analyzed both UKB and CKB protein data together in the combined training dataset without the use of bridging samples, which were not available. It is likely that there may be differences in normalization between the UKB and CKB, but the protein expression ranges were remarkably similar between the two cohorts. Third, our datasets do not have DNAm data that would allow for direct comparisons between proteomic and DNAm clocks.

In summary, our analyses provide strong evidence that plasma proteomics are a powerful tool for measuring biological age and can be used to quantify a biological aging signature that is involved in most common age-related diseases. Moreover, our study provides an important bridge between pathways identified in basic aging research and rates of age-related disease and multimorbidity studied in clinical and population research. Further work is needed to refine quantification of proteomic aging and to elucidate the genetic and environmental determinants of accelerated proteomic aging. Our work demonstrates that development of proteomic aging clocks can be used as a robust tool to help to identify protein targets or pathways for possible drug treatment or lifestyle modifications to reduce premature and delay the onset of major age-related diseases.

## Extended Data Figures

**Extended Data Fig. 1.**
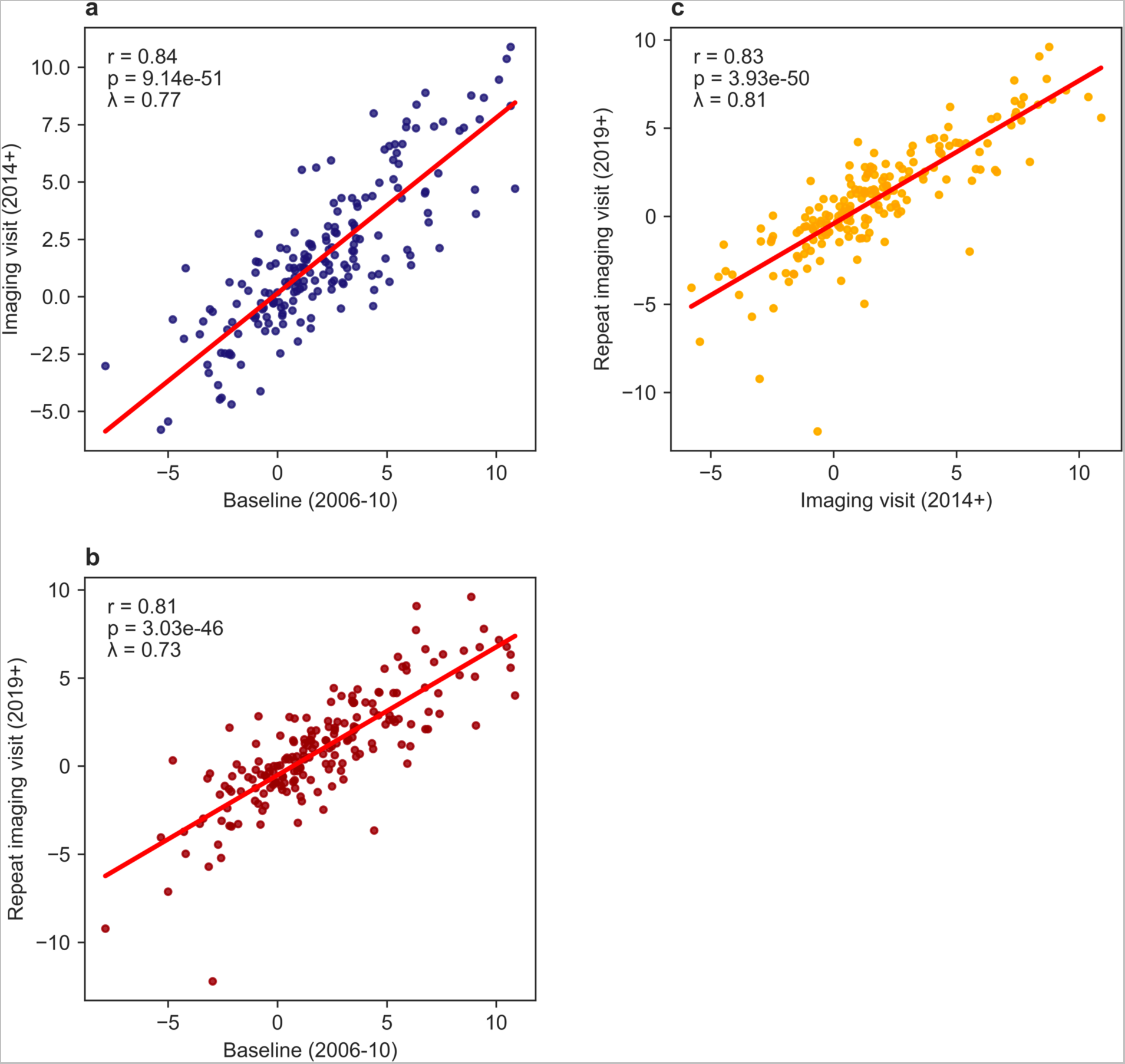
Stability of ProtAge protein associations with age across 3 time points. **a)** Comparison of betas for the association between each of the 226 ProtAge APs and age during baseline (n=45,117) and during the imaging visit (n=118). **b)** Comparison of betas for the association between each of the 226 ProtAge APs and age during baseline (n=45,117) and during the repeat imaging visit (n=108). **c)** Comparison of betas for the association between each of the 226 ProtAge APs and age during the imaging visit (n=118) and during the repeat imaging visit (n=108). Shown in each plot are the Pearson correlation coefficient (r), p-value for the correlation, and the model slope (λ). APs: aging-related proteins.

**Extended Data Fig. 2.**
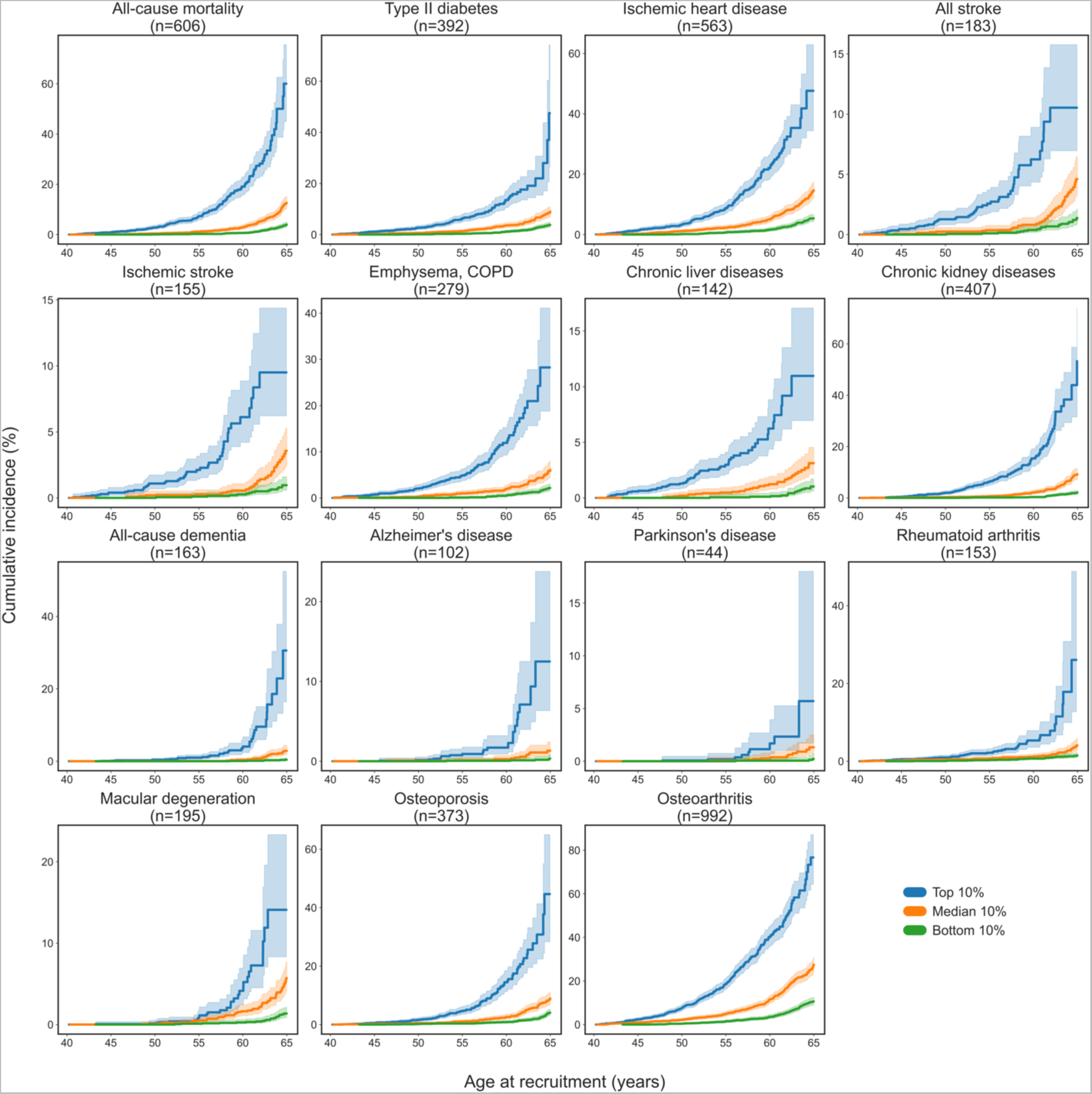
ProtAgeAccel leads to strongly diverging age-specific mortality and disease risk trajectories (women only). Cumulative incidence plots for the top, median, and bottom deciles of ProtAgeAccel among UK Biobank women (n=24,409). Number of incident cases are shown for each disease – these numbers reflect the total number of incident cases present only among those in the 3 deciles shown, not the full dataset. Incidence rates are shown for the subssequent 11-16 years of follow-up after recruitment for each given age at recruitment (e.g., the cumulative incidence rate shown at age 65 is the rate of incident cases in the 11-16 years of follow up in those aged 65 years at recruitment). All plots show 95% confidence intervals in lighter shading. ProtAgeAccel: proteomic age acceleration (in years).

**Extended Data Fig. 3.**
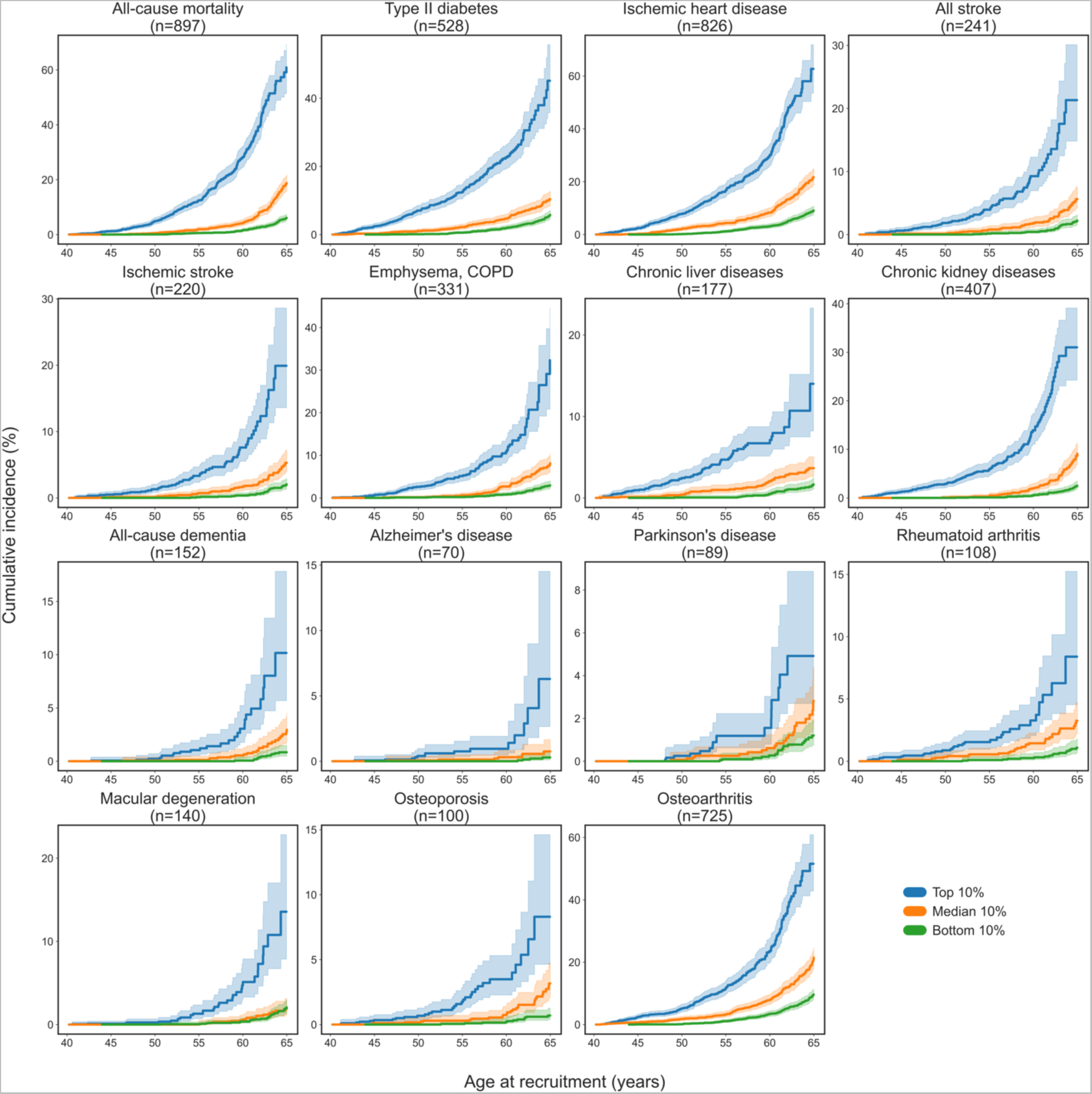
ProtAgeAccel leads to strongly diverging age-specific mortality and disease risk trajectories (men only). Cumulative incidence plots for the top, median, and bottom deciles of ProtAgeAccel among UK Biobank men (n=20,708). Number of incident cases are shown for each disease – these numbers reflect the total number of incident cases present only among those in the 3 deciles shown, not the full dataset. Incidence rates are shown for the subssequent 11-16 years of follow-up after recruitment for each given age at recruitment (e.g., the cumulative incidence rate shown at age 65 is the rate of incident cases in the 11-16 years of follow up in those aged 65 years at recruitment). All plots show 95% confidence intervals in lighter shading. ProtAgeAccel: proteomic age acceleration (in years).

**Extended Data Fig. 4.**
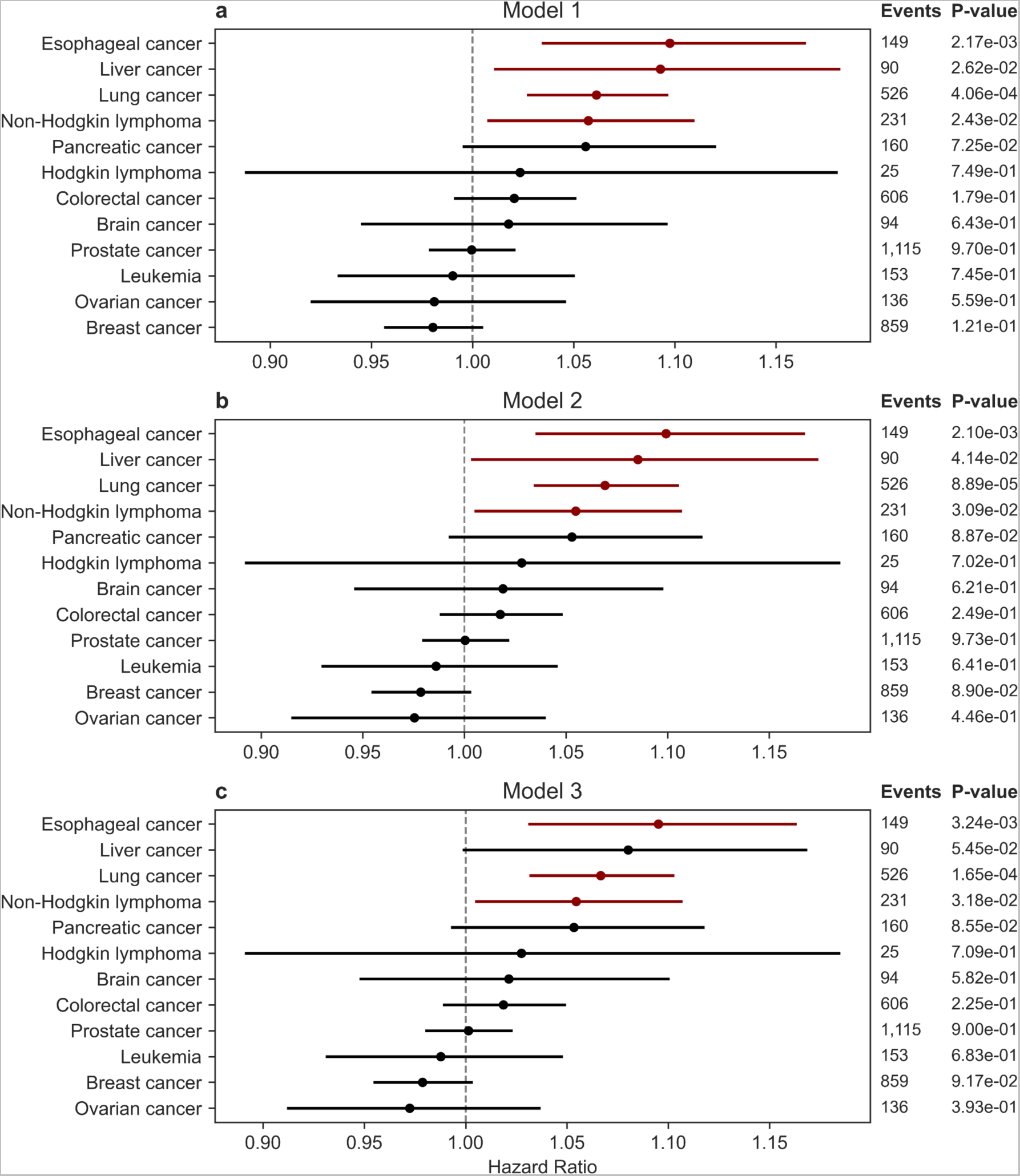
Associations between ProtAgeAccel and cancers in the UKB. Associations between ProtAgeAccel and and incident cancer diagnoses in Cox proportional hazards models with increasing levels of covariate adjustment. All models were run in the UK Biobank (UKB; n=45,117). **a)**. Model 1 is adjusted for age and sex. **b)** Model 2 is adjusted for age, sex, Townsend deprivation index, recruitment centre, IPAQ activity group, and smoking status. **c)** Model 3 is adjusted for age, sex, Townsend deprivation index, recruitment centre, IPAQ activity group, smoking status, BMI, and prevalent hypertension. ProtAgeAccel: proteomic age acceleration (in years).

**Extended Data Fig. 5.**
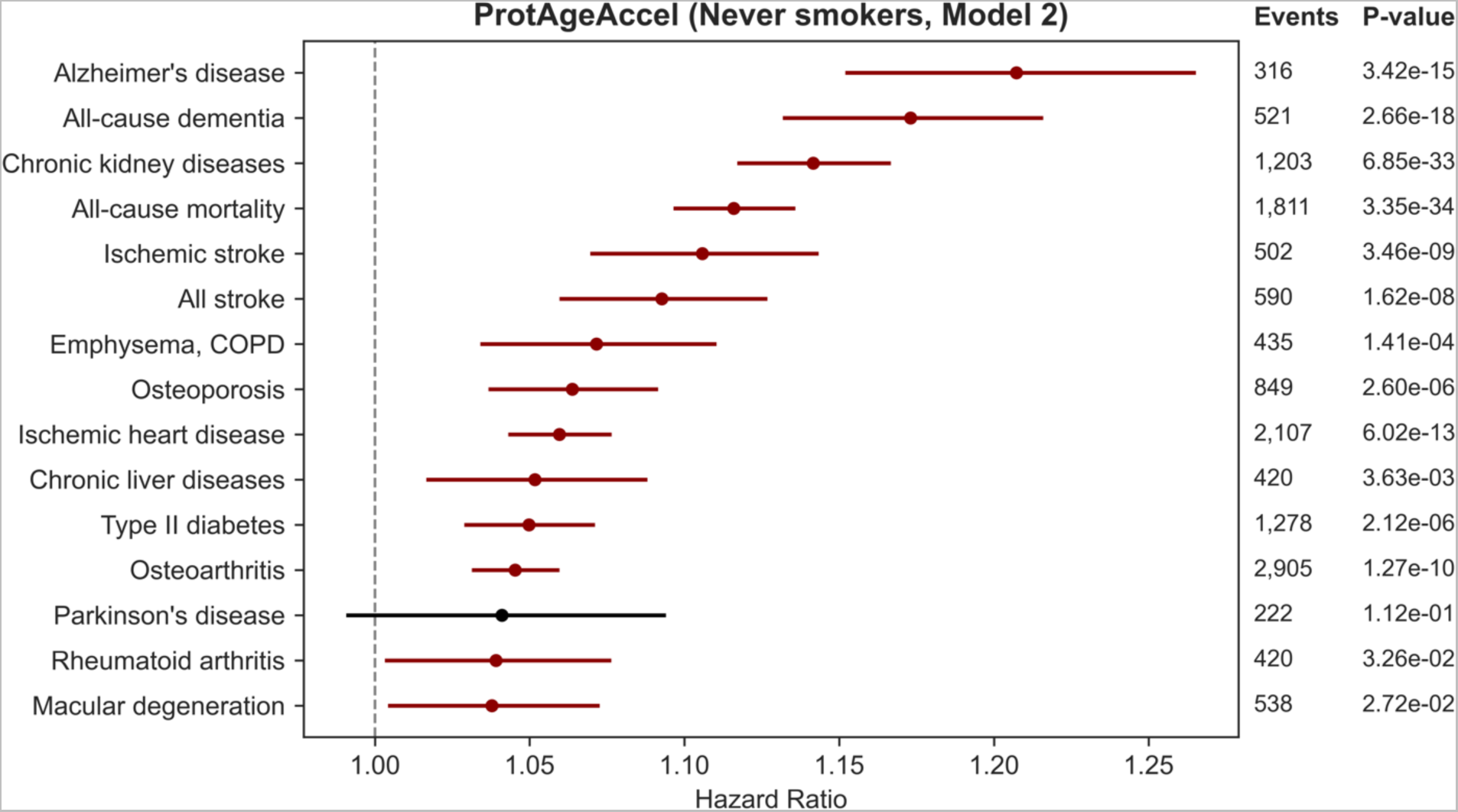
Effect size of ProtAgeAccel on mortality and disease among non-smokers. Associations between ProtAgeAccel and mortality or diseases in Cox proportional hazards models using model 2 (adjusted for age, sex, Townsend deprivation index, recruitment centre, and IPAQ activity group). The sample was subset to only UK Biobank participants who report being never smokers (n=24,360). ProtAgeAccel: proteomic age acceleration (in years).

**Extended Data Fig. 6.**
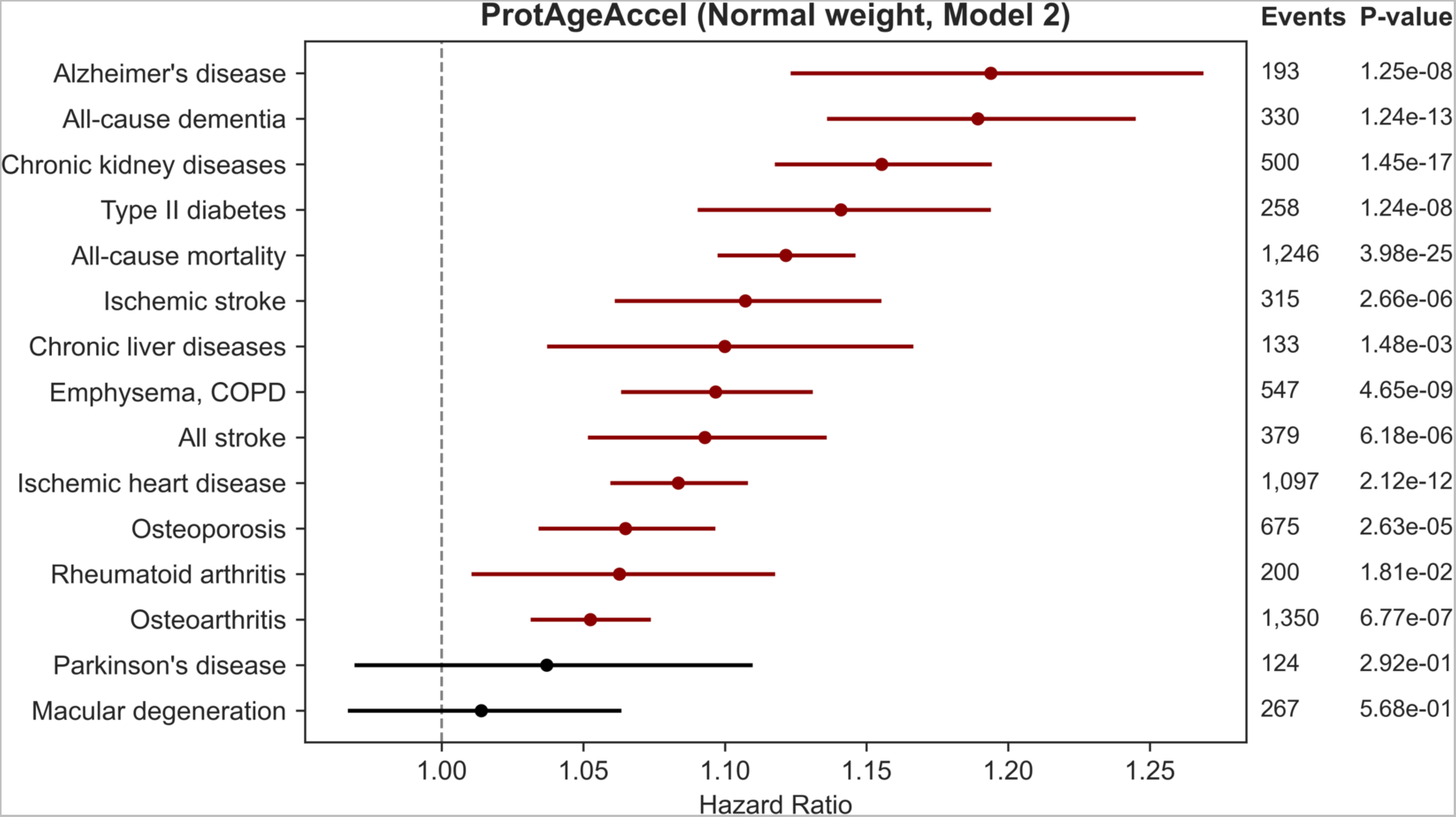
Effect size of ProtAgeAccel on mortality and disease among those in normal weight range. Associations between ProtAgeAccel and and mortality or diseases in Cox proportional hazards models using model 2 (adjusted for age, sex, Townsend deprivation index, recruitment centre, and IPAQ activity group). The sample was subset to only UK Biobank participants with a BMI ≥ 18.5 and BMI < 25 (n=14,453). ProtAgeAccel: proteomic age acceleration (in years).

**Extended Data Fig. 7.**
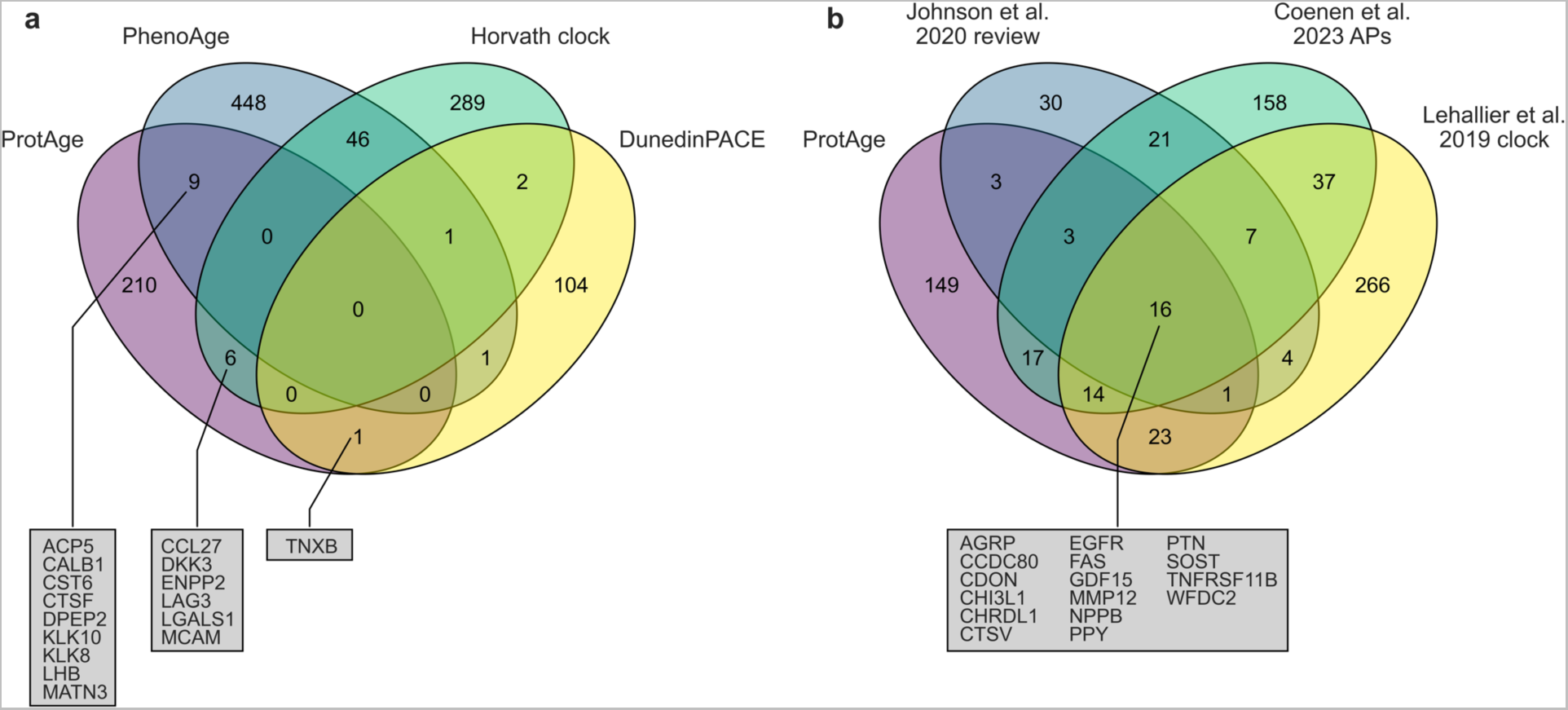
Overlap of ProtAge APs with existing DNAm and proteomic clock publications. **a)** Overlap between genes coding for the 226 ProtAge APs versus genes mapped by proximity to CpGs from common DNAm clocks. **b)** Overlap between 226 ProtAge APs versus a recent systematic review of APs (Johnson et al. 2020), a recent comprehensive analysis of SOMAscan proteins associated with age (Coenen et al. 2023), and a recent proteomic aging clock created using SOMAscan data (Lehallier et al. 2019). APs: aging-related proteins, DNAm: DNA methylation.

## Extended Data Tables

**Extended Data Table 1.**
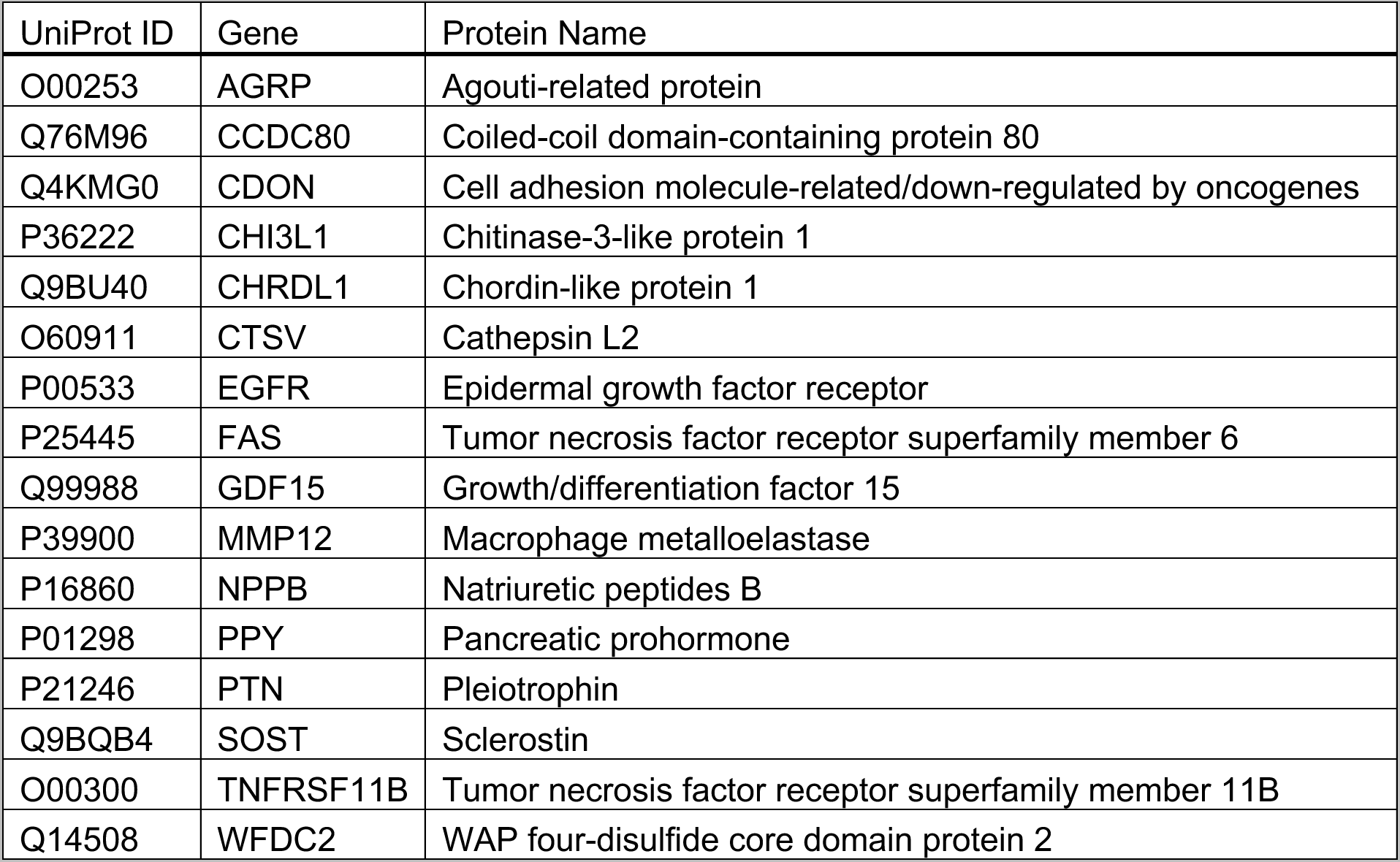
ProtAge APs also associated with age in other major publications. Proteins listed are those also reported to associate with age in Johnson et al. (2020), Coenen et al. (2023), and Lehallier et al. (2019). APs: aging-related proteins.

## Methods

### Study participants

The UK Biobank (UKB) is a prospective cohort study with extensive genetic and phenotype data available for 502,505 individuals resident in the United Kingdom who were recruited from 2006-2010.^20^ The full [UK Biobank protocol](https://www.ukbiobank.ac.uk/media/gnkeyh2q/study-rationale.pdf) is available online. We restricted our UKB sample to those participants with Olink Explore 1536 data available at baseline who were randomly sampled from the main UKB population (n=45,117).

The China Kadoorie Biobank (CKB) is a prospective cohort study of 512,724 adults aged 30-79 years who were recruited from ten geographically diverse (five rural and five urban) areas across China during 2004-2008. Details on the CKB study design and methods have been previously reported.^21^ We restricted our CKB sample to those participants with Olink Explore data available at baseline who were randomly sampled from the main CKB population, and who were genetically unrelated to each other and had no prior history of cardiovascular disease or statin use at baseline (n=2,026).

### Proteomic profiling

Proteomic profiling in the UKB and CKB was carried out for protein analytes measured via the Olink Explore 1536 platform that links four Olink panels (Cardiometabolic, Inflammation, Neurology, and Oncology). The random subsample of UKB proteomics participants (n=45,117) were selected by removing those in batches 0 and 7. Randomized participants selected for proteomic profiling in the UKB have been shown previously to be highly representative of the wider UKB population.^22^ UKB Olink data are provided Normalized Protein eXpression (NPX) values on a log2 scale, with details on sample selection, processing, quality control, and normalization [documented online](https://biobank.ndph.ox.ac.uk/showcase/label.cgi?id=1839).

In the CKB, stored baseline plasma samples from participants were retrieved, thawed, and sub-aliquoted to multiple aliquots, with one (100 µL) shipped on dry ice to the Olink Bioscience Laboratory at Uppsala, Sweden, for proteomic analysis using a multiplex proximity extension assay. To minimize inter- and intra-run variation, the samples were randomized across plates and normalized using both an internal control (extension control) and an inter-plate control and then transformed using a pre-determined correction factor. The limit of detection (LOD) was determined using negative control samples (buffer without antigen). A sample was flagged as having QC warning if the incubation control deviated more than a pre-determined value (±0.3) from the median value of all samples on the plate (but values below LOD were included in the analyses). The pre-processed data were provided in the arbitrary NPX unit on a log2 scale.

We excluded three proteins from analysis that were missing in over 10% of the UKB sample (CTSS, PCOLCE, NPM1), leaving a total of 1,459 proteins for analysis. Protein expression values were not imputed, as the LightGBM model used can handle missing values.

### Outcomes

UKB aging biomarkers were measured using baseline non-fasting blood serum samples as previously described.^23^ Biomarkers were previously adjusted for technical variation by the UKB, with [sample processing](https://biobank.ndph.ox.ac.uk/showcase/showcase/docs/serum_biochemistry.pdf) and [quality control](https://biobank.ndph.ox.ac.uk/showcase/ukb/docs/biomarker_issues.pdf) procedures described on the UK Biobank website. Field IDs for all biomarkers and measures of physical and cognitive decline are shown in Table S3. Poor self-rated health, slow walking pace, self-rated facial aging, feeling tired/lethargic every day, and frequent insomnia were all binary dummy variables coded as all other responses versus responses for “Poor” (overall health rating; Field ID 2178), “Slow pace” (usual walking pace; Field ID 924), “Older than you are” (facial aging; Field ID 1757), “Nearly every day” (frequency of tiredness / lethargy in last 2 weeks; Field ID 2080), and “Usually” (sleeplessness / insomnia; Field ID 1200), respectively. Sleeping 10+ hours/day was coded as a binary variable using the continuous measure of self-reported sleep duration (Field ID 160). Systolic and diastolic blood pressure were averaged across both automated readings. Standardized lung function (FEV1) was calculated by dividing the FEV1 best measure (field ID 20150) by standing height squared (field ID 50). Hand grip strength variables (field ID 46,47) were divided by weight (Field ID 21002) to normalize according to body mass. Frailty index was calculated using the algorithm previously developed for UK Biobank data by Williams et al. (2019).^24^ Components of the frailty index are shown in Table S4.

Detailed information about the [linkage procedure](http://biobank.ctsu.ox.ac.uk/crystal/refer.cgi?id=115559) with national registries for mortality and cause of death information in the UKB is available online. Mortality data were accessed from the UKB data portal on May 23, 2023, with a censoring date of November 30, 2022 for all participants (12-16 years of follow-up).

Data used to define prevalent and incident chronic diseases in the UKB are outlined in Table S5. In the UKB, incident cancer diagnoses were ascertained using ICD diagnosis codes and corresponding dates of diagnosis from linked cancer and mortality register data. Incident diagnoses for all other diseases were ascertained using ICD diagnosis codes and corresponding dates of diagnosis taken from linked hospital inpatient, primary care, and death register data. Primary care read codes were converted to corresponding ICD diagnosis codes using the [lookup table](https://biobank.ndph.ox.ac.uk/ukb/refer.cgi?id=592) provided by the UKB. Linked hospital inpatient, primary care, and cancer register data were accessed from the UKB data portal on May 23, 2023, with a censoring date of October 31, 2022; July 31, 2021; or February 28, 2018 for participants recruited in England, Scotland, or Wales, respectively (8-16 years of follow-up).

In the CKB, information about incident disease and cause-specific mortality was obtained by electronic linkage, via the unique national identification number, to established local mortality (cause-specific) and morbidity (for stroke, IHD, cancer and diabetes) registries and to the health insurance system that records any hospitalization episodes and procedures.^21,25^ All disease diagnoses were coded using the Tenth International Classification of Diseases (ICD-10), blinded to any baseline information and participants were followed up to death, loss-to-follow-up or the 1 January 2019. ICD-10 codes used to define diseases studied in the CKB are shown in Table S6.

### Missing data imputation

Missing values for all UKB data except for Olink expression data, age, and incident health outcomes were imputed using the R package missRanger,^26^ which combines random forest imputation with predictive mean matching. We imputed a single dataset using a maximum of 10 iterations and 200 trees. All other random forest hyperparameters were left at their default. The imputation dataset included all baseline variables available in the UKB as predictors for imputation, excluding variables with any nested response patterns. Responses of “do not know” were set to NA and imputed. Responses of “prefer not to answer” were not imputed and set to NA in the final analysis dataset. CKB data had no missing values to impute.

### Calculation of chronological age measures

We calculated ourselves a chronological age at recruitment variable as a decimal in the UKB, since age at recruitment (field ID 21022) is only provided as a whole integer value. This was done by taking month of birth (field ID 52) and year of birth (field ID 34) and creating an approximate date of birth for each participant as the first day of their birth month and year. Age at recruitment as a decimal value was then calculated as the number of days between each participant’s recruitment date (field ID 53) and approximate birth date divided by 365.25. Age at the first imaging follow-up (2014+) and the repeat imaging follow-up (2019+) were then calculated by taking the number of days between the date of each participant’s follow-up visit and their initial recruitment date divided by 365.25 and adding this to age at recruitment as a decimal value. Recruitment age in the CKB is already provided as a decimal value.

### Calculation of ProtAge

UKB and CKB data were both split into 70/30 train/test splits. Using the combined train data from both cohorts (n=32,999), we trained a model to predict age at recruitment using all 1,459 proteins in a single LightGBM^27^ model. First, model hyperparameters were tuned via 5-fold cross-validation using the Optuna module in Python,^28^ with parameters tested across 300 trials and optimized to maximize the average R^2^ of the models across all folds. We then carried out Boruta feature selection via the shap-hypetune module. Boruta is an algorithm developed to select all relevant features in a prediction model by comparing the performance of real features to randomly permuted shadow features.^29^ When running Boruta, we used 200 trials and a threshold of 100% to compare shadow and real features (meaning that a real feature is selected if it performs better than all shadow features). Third, we re-tuned model hyperparameters for a new model with the subset of selected proteins using the same procedure as before. Both tuned LightGBM models before and after feature selection were checked for overfitting and validated by performing 5-fold cross-validation in the combined train set and testing the performance of the model against the combined holdout test sets from both cohorts, as well as individually using the holdout test set from each cohort separately. Across all analysis steps, LightGBM models were run with 5,000 estimators, 20 early stopping rounds, and using R^2^ as a custom evaluation metric to identify the model that explained the maximum variation in age (according to R^2^).

Once the final model was trained and validated, we calculated protein predicted age (ProtAge) for the entire combined sample of both cohorts (n=47,143) using 5-fold cross-validation. Within each fold, a LightGBM model was trained using the final hyperparameters and predicted age values were generated for the test set of that fold. We then combined the predicted age values from each of the folds to create a measure of protein predicted age (ProtAge) for the entire sample. ProtAge was then mapped to the participant IDs in each cohort and separated into each cohort dataset. Finally, we calculated proteomic aging acceleration (ProtAgeAccel) by taking the difference of ProtAge minus chronological age at recruitment separately in each cohort.

### Statistical analysis

All statistical analyses were carried out using Python v.3.6 and R v.4.2.2. All associations between ProtAgeAccel and aging biomarkers and physical/cognitive decline measures in the UKB were tested using linear/logistic regression using the statsmodels module.^30^ All models were adjusted for age, sex, Townsend deprivation index, assessment center, self-reported ethnicity (Black, white, Asian, Mixed, Other), IPAQ activity group (low, moderate, high), and smoking status (never, previous, current). P-values were not corrected for multiple comparisons.

All associations between ProtAgeAccel and incident outcomes (mortality, 26 diseases) were tested using Cox proportional hazards models using the lifelines module.^31^ Survival outcomes were defined using follow-up time to event and the binary incident event indicator. For all incident disease outcomes, prevalent cases were excluded from the dataset before models were run. For all incident outcome Cox modelling in the UKB, three successive models were tested with increasing numbers of covariates. Model 1 included adjustment for age at recruitment and sex. Model 2 included all model 1 covariates, plus Townsend deprivation index (Field ID 22189), assessment center (Field ID 54), physical activity (IPAQ activity group; Field ID 22032), and smoking status (Field ID 20116). Model 3 included all model 3 covariates plus BMI (Field ID 21001) and prevalent hypertension (definition in Table S5). P-values were not corrected for multiple comparisons.

Functional enrichments (GO biological processes, GO molecular function, KEGG, Reactome) and protein-protein interaction (PPI) networks were downloaded from STRING (v.11.5) using the STRING API in Python. Since the Olink Explore panel consists of a subset of proteins selected to be involved in disease, it represents a possibly biased background and we did not use the subset of Olink Explore proteins as a custom background for enrichment analyses. We only considered PPIs from STRING at a high level of confidence (>0.7) from the co-expression data.

SHAP interaction values from the trained LightGBM ProtAge model were retrieved using the shap module.^32,33^ SHAP-based PPI networks were generated by first taking the average of the absolute value of each protein-protein SHAP interaction score across all samples. We then used an interaction threshold of 0.009 and removed all interactions below this threshold, which yielded a subset of variables similar in number to the node degree > 2 threshold used for the STRING PPI network. Both SHAP-based and STRING^34^-based PPI networks were visualized and plotted using the NetworkX module.^35^

Cumulative incidence curves and survival tables for deciles of ProtAgeAccel were calculated using KaplanMeierFitter from the lifelines module. Since our data were right-censored, we plotted cumulative events against age at recruitment on the x-axis. All plots were generated using matplotlib^36^ and seaborn.^37^

## Data Availability

UK Biobank data are available through a procedure described at: https://www.ukbiobank.ac.uk/enable-your-research. The China Kadoorie Biobank (CKB) is a global resource for the investigation of lifestyle, environmental, blood biochemical and genetic factors as determinants of common diseases. The CKB study group is committed to making the cohort data available to the scientific community in China, the UK, and worldwide to advance knowledge about the causes, prevention and treatment of disease. For detailed information on what data is currently available to open access users and how to apply for it, please visit: https://www.ckbiobank.org/data-access. A research proposal will be requested to ensure that any analysis is performed by bona fide researchers. Researchers who are interested in obtaining additional information or data that underlines this paper should contact. For any data that is not currently available to open access, researchers may need to develop formal collaboration with the CKB study group.
R and Python code needed to reproduce all analyses, figures, and tables will be made publicly available on GitHub before journal publication.

https://www.ckbiobank.org/data-access

## Acknowledgements

We would like to acknowledge the UK Biobank and China Kadoorie Biobank participants for their dedication to participating in ongoing research and electronic health record linkage. All UK Biobank data was accessed under UK Biobank Application #61054. We further acknowledge the CKB project staff and the China CDC and its regional offices for assisting with CKB fieldwork. We thank Judith Mackay in Hong Kong; Yu Wang, Gonghuan Yang, Zhengfu Qiang, Lin Feng, Maigeng Zhou, Wenhua Zhao, and Yan Zhang in the China CDC; Lingzhi Kong, Xiucheng Yu, and Kun Li in the Chinese Ministry of Health; and Sarah Clark, Martin Radley, and Mike Hill in the CTSU, Oxford, for assisting with the planning, conduct and organization of the CKB study. Finally, we thank Peter Block for his clinical input on liver enzymes and biomarkers.

## Ethics approval

UK Biobank data use (Project Application Number 61054) was approved by the UK Biobank according to their established access procedures. UK Biobank has approval from the North West Multi-centre Research Ethics Committee (MREC) as a Research Tissue Bank (RTB), and as such researchers using UK Biobank data do not require separate ethical clearance and can operate under the RTB approval. The China Kadoorie Biobank (CKB) complies with all the required ethical standards for medical research on human subjects. Ethical approvals were granted and have been maintained by the relevant institutional ethical research committees in the UK and China.

## Author contributions

M.A.A., C.M.vD., N.A., S.X., D.B., and Z.C conceptualized the study. M.A.A. performed all analysis and data visualization, as well as all UKB data curation. Additional CKB data curation was performed by P.Y., M.M., and D.B., and collection of CKB data was facilitated by L.L., J.L., and Z.C. Data curation and formal analyses were supervised by C.M.vD., N.A., D.B., and Z.C. Analytical and interpretational input and was provided by L.W., A.N-H., P.Y., M.M., and R.C. for all analyses. M.A.A. prepared the manuscript, figures, tables, and supplementary files, with edits and revisions provided by all other co-authors. The GitHub code repository was created and is maintained by M.A.A.

## Data Access Statement

UK Biobank data are available through a procedure described at: https://www.ukbiobank.ac.uk/enable-your-research. The China Kadoorie Biobank (CKB) is a global resource for the investigation of lifestyle, environmental, blood biochemical and genetic factors as determinants of common diseases. The CKB study group is committed to making the cohort data available to the scientific community in China, the UK, and worldwide to advance knowledge about the causes, prevention and treatment of disease. For detailed information on what data is currently available to open access users and how to apply for it, please visit: https://www.ckbiobank.org/data-access. A research proposal will be requested to ensure that any analysis is performed by *bona fide* researchers. Researchers who are interested in obtaining additional information or data that underlines this paper should contact. For any data that is not currently available to open access, researchers may need to develop formal collaboration with the CKB study group.

R and Python code needed to reproduce all analyses, figures, and tables will be made publicly available on GitHub before journal publication.

## References

1 Niccoli, T. & Partridge, L. Ageing as a risk factor for disease. Curr Biol 22, R741–752 (2012). 10.1016/j.cub.2012.07.024

2 Partridge, L., Deelen, J. & Slagboom, P. E. Facing up to the global challenges of ageing. Nature 561, 45–56 (2018). 10.1038/s41586-018-0457-8

3 Chang, A. Y., Skirbekk, V. F., Tyrovolas, S., Kassebaum, N. J. & Dieleman, J. L. Measuring population ageing: an analysis of the Global Burden of Disease Study 2017. Lancet Public Health 4, e159–e167 (2019). 10.1016/S2468-2667(19)30019-2

4 Langenberg, C., Hingorani, A. D. & Whitty, C. J. M. Biological and functional multimorbidity-from mechanisms to management. Nat Med 29, 1649–1657 (2023). 10.1038/s41591-023-02420-6

5 Horvath, S. & Raj, K. DNA methylation-based biomarkers and the epigenetic clock theory of ageing. Nat Rev Genet 19, 371–384 (2018). 10.1038/s41576-018-0004-3

6 Rutledge, J., Oh, H. & Wyss-Coray, T. Measuring biological age using omics data. Nat Rev Genet 23, 715–727 (2022). 10.1038/s41576-022-00511-7

7 López-Otín, C., Blasco, M. A., Partridge, L., Serrano, M. & Kroemer, G. Hallmarks of aging: An expanding universe. Cell 186, 243–278 (2023). 10.1016/j.cell.2022.11.001

8 Coenen, L., Lehallier, B., de Vries, H. E. & Middeldorp, J. Markers of aging: Unsupervised integrated analyses of the human plasma proteome. Front Aging 4, 1112109 (2023). 10.3389/fragi.2023.1112109

9 Johnson, A. A., Shokhirev, M. N., Wyss-Coray, T. & Lehallier, B. Systematic review and analysis of human proteomics aging studies unveils a novel proteomic aging clock and identifies key processes that change with age. Ageing Res Rev 60, 101070 (2020). 10.1016/j.arr.2020.101070

10 Lehallier, B. et al. Undulating changes in human plasma proteome profiles across the lifespan. Nat Med 25, 1843–1850 (2019). 10.1038/s41591-019-0673-2

11 Lehallier, B., Shokhirev, M. N., Wyss-Coray, T. & Johnson, A. A. Data mining of human plasma proteins generates a multitude of highly predictive aging clocks that reflect different aspects of aging. Aging Cell 19, e13256 (2020). 10.1111/acel.13256

12 Kabacik, S. et al. The relationship between epigenetic age and the hallmarks of aging in human cells. Nat Aging 2, 484–493 (2022). 10.1038/s43587-022-00220-0

13 Levine, M. E. et al. An epigenetic biomarker of aging for lifespan and healthspan. Aging (Albany NY) 10, 573–591 (2018). 10.18632/aging.101414

14 Lu, A. T. et al. DNA methylation GrimAge strongly predicts lifespan and healthspan. Aging (Albany NY) 11, 303–327 (2019). 10.18632/aging.101684

15 Belsky, D. W. et al. DunedinPACE, a DNA methylation biomarker of the pace of aging. Elife 11 (2022). 10.7554/eLife.73420

16 Horvath, S. DNA methylation age of human tissues and cell types. Genome Biol 14, R115 (2013). 10.1186/gb-2013-14-10-r115

17 Pietzner, M. et al. Synergistic insights into human health from aptamer- and antibody-based proteomic profiling. Nat Commun 12, 6822 (2021). 10.1038/s41467-021-27164-0

18 Giannini, E. G., Testa, R. & Savarino, V. Liver enzyme alteration: a guide for clinicians. CMAJ 172, 367–379 (2005). 10.1503/cmaj.1040752

19 Aravinthan, A. et al. The senescent hepatocyte gene signature in chronic liver disease. Exp Gerontol 60, 37–45 (2014). 10.1016/j.exger.2014.09.011

20 Palmer, L. UK Biobank: bank on it. Lancet 369, 1980–1982 (2007). 10.1016/S0140-6736(07)60924-6

21 Chen, Z. et al. China Kadoorie Biobank of 0.5 million people: survey methods, baseline characteristics and long-term follow-up. Int J Epidemiol 40, 1652–1666 (2011). 10.1093/ije/dyr120

22 Sun, B. B. et al. Genetic regulation of the human plasma proteome in 54,306 UK Biobank participants. bioRxiv (2022). 10.1101/2022.06.17.496443

23 Elliott, P. & Peakman, T. C. The UK Biobank sample handling and storage protocol for the collection, processing and archiving of human blood and urine. International Journal of Epidemiology 37, 234–244 (2008). 10.1093/ije/dym276

24 Williams, D. M., Jylhävä, J., Pedersen, N. L. & Hägg, S. A Frailty Index for UK Biobank Participants. J Gerontol A Biol Sci Med Sci 74, 582–587 (2019). 10.1093/gerona/gly094

25 Chen, Z. et al. Cohort profile: the Kadoorie Study of Chronic Disease in China (KSCDC). Int J Epidemiol 34, 1243–1249 (2005). 10.1093/ije/dyi174

26 Mayer, M. *missRanger: Fast Imputation of Missing Values. R package version 2.1.0*., <https://CRAN.R-project.org/package=missRanger> (2019).

27 Ke, G. et al. LightGBM: A Highly Efficient Gradient Boosting Decision Tree. Advances in Neural Information Processing Systems 30, 3149–3157 (2017).

28 Akiba, T., Sano, S., Yanase, T., Ohta, T. & Koyama, M. Optuna: A Next-generation Hyperparameter Optimization Framework. KDD ’19: Proceedings of the 25th ACM SIGKDD International Conference on Knowledge Discovery & Data Mining, 2623–2631 (2019). 10.1145/3292500.3330701

29 Kursa, M. B., Jankowski, A. & Rudnicki, W. R. Boruta – A System for Feature Selection. Fundamenta Informaticae 101, 271–285 (2010).

30 Skipper, S. & Perktold, J. Statsmodels: Econometric and statistical modeling with python. Proceedings of the 9th Python in Science Conference (2010).

31 Davidson-Pilon, C. lifelines, survival analysis in Python. (2023). 10.5281/zenodo.7883870

32 Lundberg, S. M. et al. From Local Explanations to Global Understanding with Explainable AI for Trees. Nat Mach Intell 2, 56–67 (2020). 10.1038/s42256-019-0138-9

33 Lundberg, S. M. & Lee, S.-I. A unified approach to interpreting model predictions. Advances in Neural Information Processing Systems 30, 4765–4774 (2017).

34 Szklarczyk, D. et al. STRING v10: protein-protein interaction networks, integrated over the tree of life. Nucleic Acids Res 43, D447–452 (2015). 10.1093/nar/gku1003

35 Hagberg, A., Schult, A. & Swart, P. in Proceedings of the 7th Python in Science conference (SciPy 2008). (eds G Varoquaux, T Vaught, & J Millman) 11–15

36 Hunter, J. D. Matplotlib: A 2D Graphics Environment. Computing in Science & Engineering 9, 90–95 (2007). 10.1109/MCSE.2007.55

37 Waskom, M. L. seaborn: statistical data visualization. Journal of Open Source Software 6, 3021 (2021). 10.21105/joss.03021

