## Supplementary Information for "Proteomic aging clock predicts mortality and risk of common age-related diseases in diverse populations"

Argentieri et al. 2023

### SUPPLEMENTARY INFORMATION

### Supplementary Figures

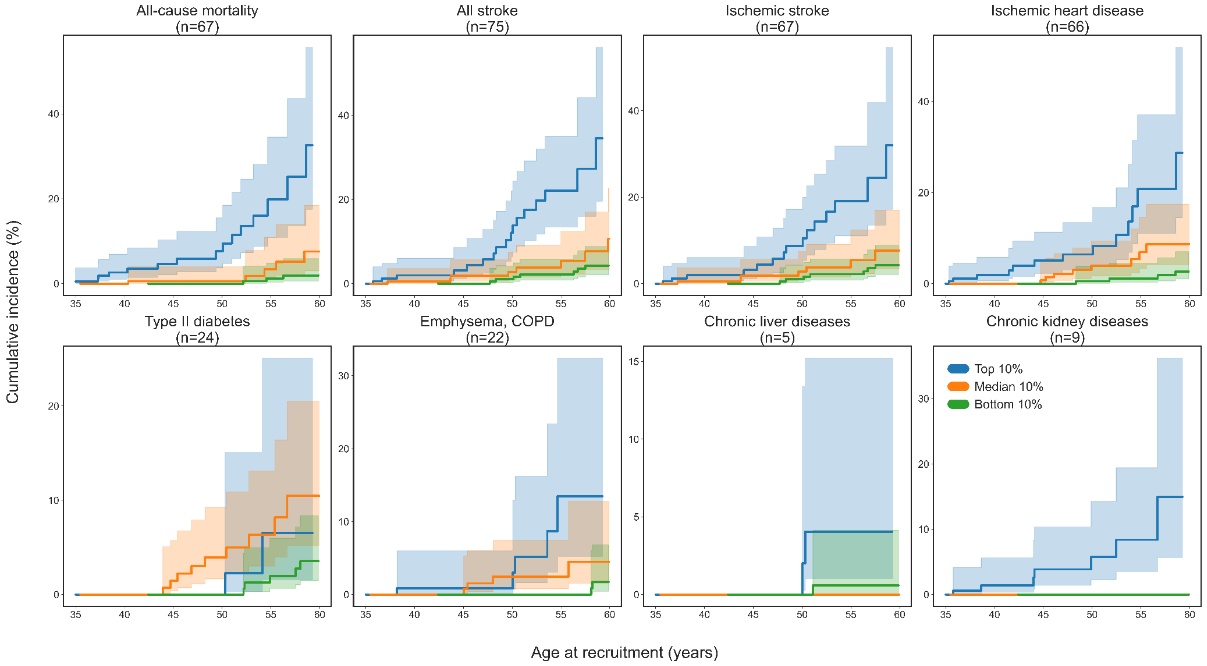

**Fig. S1. Age-specific cumulative incidence for mortality and disease risk in the CKB.** Cumulative incidence plots for the top, median, and bottom deciles of ProtAgeAccel in the China Kadoorie Biobank (CKB; n=2,026). Number of incident cases are shown for each disease – these numbers reflect the total number of incident cases present only among those in the 3 deciles shown, not the full dataset. Incidence rates are shown for the 11-14 years of follow up after recruitment for each given age at recruitment (e.g., the cumulative incidence rate shown at age 60 is the rate of incident cases in the 11-16 years of follow up after recruitment for those aged 60 years at recruitment).

### Supplementary Tables

**Table S1.** Age-specific incidence rates in the UK Biobank for mortality and age-related diseases by ProtAgeAccel deciles

| Outcome | ProtAgeAccel decile | 50 years | 55 years | 60 years | 65 years |
|---|---|---|---|---|---|
| All-cause mortality | Top 10% | 2.78 | 7.34 | 19.07 | 60.02 |
| All-cause mortality | Median 10% | 0.43 | 1.11 | 2.87 | 12.60 |
| All-cause mortality | Bottom 10% | 0.05 | 0.24 | 0.62 | 3.99 |
| Type II diabetes | Top 10% | 2.67 | 6.33 | 13.47 | 47.49 |
| Type II diabetes | Median 10% | 0.62 | 1.30 | 3.53 | 8.99 |
| Type II diabetes | Bottom 10% | 0.10 | 0.30 | 1.14 | 3.75 |
| Ischemic heart disease | Top 10% | 3.26 | 8.76 | 22.04 | 47.60 |
| Ischemic heart disease | Median 10% | 1.12 | 2.28 | 5.02 | 14.65 |
| Ischemic heart disease | Bottom 10% | 0.16 | 0.67 | 1.58 | 5.34 |
| All stroke | Top 10% | 1.27 | 2.57 | 6.24 | 10.53 |
| All stroke | Median 10% | 0.24 | 0.36 | 0.81 | 4.60 |
| All stroke | Bottom 10% | 0.00 | 0.10 | 0.37 | 1.38 |
| Ischemic stroke | Top 10% | 1.09 | 2.12 | 6.12 | 9.50 |
| Ischemic stroke | Median 10% | 0.19 | 0.26 | 0.55 | 3.57 |
| Ischemic stroke | Bottom 10% | 0.00 | 0.10 | 0.26 | 0.96 |
| Emphysema, COPD | Top 10% | 2.02 | 4.87 | 11.91 | 28.23 |
| Emphysema, COPD | Median 10% | 0.24 | 0.99 | 1.92 | 6.08 |
| Emphysema, COPD | Bottom 10% | 0.00 | 0.05 | 0.50 | 2.15 |
| Chronic liver diseases | Top 10% | 1.29 | 2.97 | 6.23 | 10.96 |
| Chronic liver diseases | Median 10% | 0.20 | 0.48 | 1.23 | 3.12 |
| Chronic liver diseases | Bottom 10% | 0.00 | 0.05 | 0.10 | 1.02 |
| Chronic kidney diseases | Top 10% | 1.91 | 6.27 | 15.36 | 53.27 |
| Chronic kidney diseases | Median 10% | 0.28 | 0.63 | 2.09 | 9.21 |
| Chronic kidney diseases | Bottom 10% | 0.00 | 0.15 | 0.32 | 2.10 |
| All-cause dementia | Top 10% | 0.37 | 0.99 | 4.04 | 30.57 |
| All-cause dementia | Median 10% | 0.05 | 0.05 | 0.36 | 2.84 |
| All-cause dementia | Bottom 10% | 0.00 | 0.00 | 0.05 | 0.41 |
| Alzheimer's disease | Top 10% | 0.13 | 0.90 | 1.70 | 12.49 |
| Alzheimer's disease | Median 10% | 0.05 | 0.11 | 0.26 | 1.32 |
| Alzheimer's disease | Bottom 10% | 0.00 | 0.05 | 0.05 | 0.35 |
| Parkinson's disease | Top 10% | 0.07 | 0.18 | 1.68 | 5.70 |
| Parkinson's disease | Median 10% | 0.00 | 0.06 | 0.28 | 1.32 |
| Parkinson's disease | Bottom 10% | 0.00 | 0.00 | 0.05 | 0.22 |
| Rheumatoid arthritis | Top 10% | 0.94 | 2.17 | 5.33 | 26.06 |
| Rheumatoid arthritis | Median 10% | 0.41 | 0.71 | 1.14 | 4.09 |
| Rheumatoid arthritis | Bottom 10% | 0.05 | 0.30 | 0.68 | 1.47 |
| Macular degeneration | Top 10% | 0.12 | 0.82 | 4.14 | 14.09 |

| | | | | | |
|---|---|---|---|---|---|
| Macular degeneration | Median 10% | 0.05 | 0.51 | 1.63 | 5.69 |
| Macular degeneration | Bottom 10% | 0.00 | 0.10 | 0.26 | 1.35 |
| Osteoporosis | Top 10% | 1.58 | 4.58 | 14.48 | 44.63 |
| Osteoporosis | Median 10% | 0.48 | 1.03 | 2.50 | 8.93 |
| Osteoporosis | Bottom 10% | 0.20 | 0.35 | 0.80 | 4.04 |
| Osteoarthritis | Top 10% | 7.58 | 18.69 | 40.15 | 76.65 |
| Osteoarthritis | Median 10% | 2.21 | 4.92 | 11.53 | 27.47 |
| Osteoarthritis | Bottom 10% | 0.41 | 1.49 | 3.51 | 10.63 |

Cumulative incidence rates are shown for those who are aged 50, 55, 60, and 65 years at recruitment in the UK Biobank (n=45,117). Incidence rates are for the 11-16 years after recruitment in the UK Biobank.

| **Table S2.** Age-specific incidence rates in the China Kadoorie Biobank for mortality and age-related diseases by ProtAgeAccel deciles | | | | | | | | |
|---|---|---|---|---|---|---|---|---|
| **Outcome** | **ProtAgeAccel decile** | **35 years** | **40 years** | **45 years** | **50 years** | **55 years** | **60 years** | **65 years** |
| All-cause mortality | Top 10% | 0.53 | 2.64 | 4.65 | 7.63 | 19.82 | 32.65 | 32.65 |
| All-cause mortality | Median 10% | 0.00 | 0.00 | 0.57 | 0.57 | 3.39 | 7.57 | 7.57 |
| All-cause mortality | Bottom 10% | 0.00 | 0.00 | 0.00 | 0.00 | 1.24 | 1.93 | 4.94 |
| All stroke | Top 10% | 0.00 | 1.97 | 3.17 | 12.09 | 22.09 | 34.55 | 47.64 |
| All stroke | Median 10% | 0.00 | 0.52 | 1.85 | 2.78 | 5.42 | 10.65 | 18.74 |
| All stroke | Bottom 10% | 0.00 | 0.00 | 0.00 | 1.06 | 2.18 | 4.29 | 11.00 |
| Ischemic stroke | Top 10% | 0.00 | 1.97 | 3.17 | 8.67 | 19.06 | 32.01 | 45.61 |
| Ischemic stroke | Median 10% | 0.00 | 0.52 | 1.85 | 2.78 | 5.42 | 7.67 | 16.03 |
| Ischemic stroke | Bottom 10% | 0.00 | 0.00 | 0.00 | 1.06 | 2.18 | 4.29 | 8.94 |
| Ischemic heart disease | Top 10% | 0.00 | 1.89 | 5.09 | 6.41 | 20.77 | 28.69 | 28.69 |
| Ischemic heart disease | Median 10% | 0.00 | 0.00 | 0.70 | 3.96 | 6.95 | 8.70 | 27.56 |
| Ischemic heart disease | Bottom 10% | 0.00 | 0.00 | 0.00 | 0.54 | 1.13 | 2.66 | 11.68 |
| Type II diabetes | Top 10% | 0.00 | 0.00 | 0.00 | 0.00 | 6.52 | 6.52 | 6.52 |
| Type II diabetes | Median 10% | 0.00 | 0.00 | 1.47 | 3.93 | 6.34 | 10.47 | 14.74 |
| Type II diabetes | Bottom 10% | 0.00 | 0.00 | 0.00 | 0.00 | 1.96 | 3.55 | 4.80 |
| Emphysema, COPD | Top 10% | 0.00 | 0.86 | 0.86 | 0.86 | 13.49 | 13.49 | 35.12 |
| Emphysema, COPD | Median 10% | 0.00 | 0.00 | 0.00 | 2.45 | 2.45 | 4.48 | 4.48 |
| Emphysema, COPD | Bottom 10% | 0.00 | 0.00 | 0.00 | 0.00 | 0.00 | 1.75 | 4.13 |
| Chronic liver diseases | Top 10% | 0.00 | 0.00 | 0.00 | 0.00 | 4.04 | 4.04 | 4.04 |
| Chronic liver diseases | Median 10% | 0.00 | 0.00 | 0.00 | 0.00 | 0.00 | 0.00 | 0.00 |
| Chronic liver diseases | Bottom 10% | 0.00 | 0.00 | 0.00 | 0.00 | 0.59 | 0.59 | 0.59 |
| Chronic kidney diseases | Top 10% | 0.00 | 1.41 | 3.86 | 5.78 | 8.40 | 14.94 | 14.94 |
| Chronic kidney diseases | Median 10% | 0.00 | 0.00 | 0.00 | 0.00 | 0.00 | 0.00 | 0.00 |
| Chronic kidney diseases | Bottom 10% | 0.00 | 0.00 | 0.00 | 0.00 | 0.00 | 0.00 | 0.00 |

Cumulative incidence rates are shown for those who are aged 35, 40, 45, 50, 55, 60, and 65 years at recruitment in the China Kadoorie Biobank (n=2,026). Incidence rates are for the 11-14 years after recruitment in the China Kadoorie Biobank.

| **Table S3.** Individual aging biomarker and frailty variables tested in the UK Biobank | |
|---|---|
| **Biomarkers** | **Field ID** |
| Alanine aminotransferase | 30620 |
| Albumin | 30600 |
| Aspartate aminotransferase | 30650 |
| High sensitivity C-reactive protein | 30710 |
| Creatinine | 30700 |
| Cystatin C | 30720 |
| Total bilirubin | 30840 |
| Gamma glutamyltransferase | 30730 |
| Insulin-like growth factor 1 (IGF-1) | 30770 |
| Leukocyte telomere length | 22192 |
| **Physical measures** | |
| Usual walking pace | 924 |
| Body mass index (BMI) | 21001 |
| Self-rated health | 2178 |
| Facial aging | 1757 |
| Hours of sleep | 1160 |
| Tiredness | 2080 |
| Insomnia | 1200 |
| Systolic blood pressure | 4080 |
| Diastolic blood pressure | 4079 |
| Arterial stiffness index | 21021 |
| Heel bone mineral density | 3148 |
| Lung function (FEV1) best measure | 20150 |
| Hand grip strength (left) | 46 |
| Hand grip strength (right) | 47 |
| **Cognitive measures** | |
| Reaction time | 20023 |
| Fluid intelligence score | 20016 |

**Table S4.** Items used to construct the frailty index in the UK Biobank

| Type of deficit | Item | Trait | Field ID | Categories | Coding in Frailty Index |
|---|---|---|---|---|---|
| *Sensory* | 1 | Glaucoma * | 20002 | no,yes | Categorized 0/1 |
| | 2 | Cataracts * | 20002 | no,yes | Categorized 0/1 |
| | 3 | Hearing difficulty | 2247 | no, yes, completely deaf | Categorized 0/1 (combined yes/deaf groups as 1) |
| *Cranial* | 4 | Migraine * | 20002 | no,yes | Categorized 0/1 |
| | 5 | Dental problems | 6149 | ulcers, painful gums, bleeding gums, loose teeth, toothache, dentures | Categorized 0/1 for none vs. any |
| *Mental wellbeing* | 6 | Self-rated health | 2178 | excellent, good, fair, poor | 0 – excellent; 0.25 – good; 0.5 - fair, 1 – poor |
| | 7 | Fatigue: frequency of tiredness / lethargy in last two weeks | 2080 | not at all, several days, more than half, nearly every day | 0, 0.25, 0.5, 1, respectively |
| | 8 | Sleep: experience of sleeplessness/insomnia | 1200 | never/rarely, sometimes, usually | Categorized 0, 0.5, 1, respectively |
| | 9 | Depressed feelings: frequency in last two weeks | 2050 | not at all, several days, more than half, nearly every day | 0 – not at all, 0.5 – several days, 0.75 - - more than half, 1 – nearly every day |
| | 10 | Self-described nervous personality | 1970 | no, yes | Categorized 0/1 |
| | 11 | Severe anxiety/ panic attacks * | 20002 | no, yes | Categorized 0/1 |
| | 12 | Common to feel loneliness | 2020 | no, yes | Categorized 0/1 |
| | 13 | Sense of misery (ever/never) | 1930 | no, yes | Categorized 0/1 |
| *Infirmity* | 14 | Infirmity: long-standing illness or disability | 2188 | no, yes | Categorized 0/1 |
| | 15 | Falls in last year | 2296 | categorical: no falls, one fall, more than one | 0, 0.5, 1, respectively |
| | 16 | Fractures/broken bones in last five years | 2463 | no, yes | Categorized 0/1 |
| *Cardiometabolic* | 17 | Diabetes * | 20002 | no, yes | Categorized 0/1 |
| | 18 | Myocardial infarction * | 20002 | no, yes | Categorized 0/1 |

| | | | | | |
|---|---|---|---|---|---|
| | 19 | Angina * | 20002 | no, yes | Categorized 0/1 |
| | 20 | Stroke * | 20002 | no, yes | Categorized 0/1 |
| | 21 | High blood pressure * | 20002 | no, yes | Categorized 0/1 |
| | 22 | Hypothyroidism * | 20002 | no, yes | Categorized 0/1 |
| | 23 | Deep-vein thrombosis * | 20002 | no, yes | Categorized 0/1 |
| | 24 | High cholesterol * | 20002 | no, yes | Categorized 0/1 |
| *Respiratory* | 25 | Breathing: wheeze in last year | 2316 | no, yes | Categorized 0/1 |
| | 26 | Pneumonia * | 20002 | no, yes | Categorized 0/1 |
| | 27 | Chronic bronchitis/emphysema * | 20002 | no, yes | Categorized 0/1 |
| | 28 | Asthma * | 20002 | no, yes | Categorized 0/1 |
| *Musculoskeletal* | 29 | Rheumatoid arthritis * | 20002 | no, yes | Categorized 0/1 |
| | 30 | Osteoarthritis * | 20002 | no, yes | Categorized 0/1 |
| | 31 | Gout * | 20002 | no, yes | Categorized 0/1 |
| | 32 | Osteoporosis * | 20002 | no, yes | Categorized 0/1 |
| *Immunological* | 33 | Hayfever, allergic rhinitis or eczema * | 20002 | no, yes | Categorized 0/1 |
| | 34 | Psoriasis * | 20002 | no, yes | Categorized 0/1 |
| *Cancer* | 35 | Any cancer diagnosis * | 2453 | no, yes | Categorized 0/1 |
| | 36 | Multiple cancers diagnosed (number reported) | 134 | Range from 0 to 6 | 0 - no cancer or single cancer, 1 - multiple cancers |
| *Pain* | 37 | Chest pain | 2335 | no, yes | Categorized 0/1 |
| | 38 | Head and/or neck pain | 6159 | no, yes (combining responses to pain in head and neck/shoulders) | Categorized 0/1 |
| | 39 | Back pain | 6159 | no, yes | Categorized 0/1 |
| | 40 | Stomach/abdominal pain | 6159 | no, yes | Categorized 0/1 |
| | 41 | Hip pain | 6159 | no, yes | Categorized 0/1 |
| | 42 | Knee pain | 6159 | no, yes | Categorized 0/1 |
| | 43 | Whole-body pain | 6159 | no, yes | Categorized 0/1 |

| | | | | | |
|---|---|---|---|---|---|
| | 44 | Facial pain | 6159 | no, yes | Categorized 0/1 |
| | 45 | Sciatica * | 20002 | no, yes | Categorized 0/1 |
| *Gastrointestinal* | 46 | Gastric reflux * | 20002 | no, yes | Categorized 0/1 |
| | 47 | Hiatus hernia * | 20002 | no, yes | Categorized 0/1 |
| | 48 | Gall stones * | 20002 | no, yes | Categorized 0/1 |
| | 49 | Diverticulitis * | 20002 | no, yes | Categorized 0/1 |

* self-reported from the baseline verbal interview. Frailty index was developed by Williams et al. 2019 in the UK Biobank.[1] To create the score, 49 items are coded using the table above. The frailty score is calculated by summing all 49 codes and dividing by the total number of items (49).

| **Table S5.** Variables used to calculate prevalence and incidence of chronic diseases and clinical risk factors in the UK Biobank | | | | |
|---|---|---|---|---|
| **Chronic diseases** | **Baseline measures (field ID)** | **Baseline verbal interview diagnosis codes** | **ICD-10 codes** | **ICD-9 codes** |
| Colorectal cancer | - | - | C18-C20 | 153, 154 |
| Lung cancer | - | - | C33, C34 | 162 |
| Esophageal cancer | - | - | C15 | 150 |
| Liver cancer | - | - | C22 | 155 |
| Pancreatic cancer | - | - | C25 | 157 |
| Brain cancer | - | - | C71 | 191 |
| Leukemia | - | - | C91-C95 | 204-208 |
| Non-Hodgkin lymphoma | - | - | C82-C86 | 200, 202 |
| Breast cancer | - | - | C50 | 174 |
| Ovarian cancer | - | - | C56, C57 | 183 |
| Prostate cancer | - | - | C61 | 185 |
| Type 2 diabetes | Taking insulin medication (6153, 6177)<br>Non-fasting blood hbA1c ≥ 48 mmol/mol (30750)<br>Non-fasting blood glucose ≥ 11.1 mmol/L (30740) | 1223 | E11 | 250 |
| Ischemic heart disease | - | 1074, 1075 | I20-I25 | 410-414 |
| Cerebrovascular diseases | - | 1081, 1086, 1491, 1583 | I60-I69 | 430-438 |
| Emphysema, COPD | - | 1112, 1472 | J43-J44 | 492 |
| Chronic liver diseases | - | 1157, 1158, 1604 | K70, K73-K74, K75.8, K76.0 | 571 |
| Chronic kidney diseases | - | 1192, 1193, 1194 | N18 | 585 |
| All-cause dementia | - | 1263 | A81.0, F00-F03, F05.1, F10.6, G30-G31, I67.3 | 331.0, 290.4, 331.1, 290.2, 290.3, 291.2, 294.1, 331.2, 331.5 |
| Vascular dementia | - | 1263 | F01, I67.3 | 290.4 |

| | | | | |
|---|---|---|---|---|
| Alzheimer’s disease | - | 1263 | F00, G30 | 331.0 |
| Parkinson’s disease and parkinsonism | - | 1262 | G20-G22 | 332 |
| Rheumatoid arthritis | - | 1464 | M05-M06 | 714 |
| Macular degeneration | - | 1528 | H35.3 | 362.5 |
| Osteoporosis | - | 1309 | M80-M81 | 733.0 |
| Osteoarthritis | - | 1465 | M15-M19 | 715 |
| **Clinical risk factors** | **Baseline measures (field ID)** | **Baseline verbal interview diagnosis codes** | **ICD-10 codes** | **ICD-9 codes** |
| Prevalent hypertension | High blood pressure diagnosis by physician (6150)<br>Taking medication for high blood pressure (6153, 6177) | 1065, 1072 | I10-I15 | 401-405 |

Verbal interview diagnosis codes are contained in the non-cancer illness (field ID 20002) variables. Incident disease case were mapped to corresponding ICD codes from the cancer register data (Field IDs 20006, 400013, 40005) and the HESIN and HESIN_DIAG data tables. For all incident diseases, additional cases were retrieve using ICD-10 codes from cause of death information from linked death register data. Baseline prevalence for all diseases and clinical risk factors was calculated for all participants using baseline measures (including verbal interview diagnosis codes) + those with an ICD diagnosis before or on the date of recruitment into the UK Biobank. Incident cases are defined as those with an ICD date of diagnosis after the date of recruitment who do not have any prevalent diagnosis. Unless specific ICD subcategories are already given with dot separators, all ICD codes listed also include all subcategories (e.g., J44 includes J44, J44.0, J44.1, J44.8, J44.9).

| Table S6. Variables used to calculate prevalence and incidence of chronic diseases and clinical risk factors in the China Kadoorie Biobank | |
|---|---|
| **Chronic diseases** | **ICD-10 codes** |
| Ischemic stroke | I63 |
| All stroke | I60-I61, I63-I64 |
| All ischemic heart disease | I20-I25 |
| Type II diabetes | E11-E14 |
| Chronic obstructive pulmonary disease | J41-J44 |
| Chronic liver disease | K70, K74-K746 |
| Chronic Kidney disease | N02-N03, N07, N11, N18) |

Unless specific ICD subcategories are already given with dot separators, all ICD codes listed also include all subcategories (e.g., J44 includes J44, J44.0, J44.1, J44.8, J44.9).

### References

1 Williams, D. M., Jylhävä, J., Pedersen, N. L. & Hägg, S. A Frailty Index for UK Biobank Participants. *J Gerontol A Biol Sci Med Sci* **74**, 582-587 (2019). https://doi.org/10.1093/gerona/gly094
